# Atlas of cardiomyopathy associated *DES* (desmin) mutations: Functional insights into the critical 1B domain

**DOI:** 10.1101/2025.09.19.25335981

**Authors:** Sabrina Voß, Hendrik Milting, Franziska Klag, Matthias Semisch, Stephanie Holler, Jonas Reckmann, Manuel Göz, Dario Anselmetti, Jan Gummert, Marcus-André Deutsch, Volker Walhorn, Andreas Brodehl

**Affiliations:** Erich and Hanna Klessmann Institute, Heart and Diabetes Center NRW, University Hospital of the Ruhr-University Bochum, Medical School OWL (University of Bielefeld), Georgstrasse 11, 32545 Bad Oeynhausen, Germany; Experimental Biophysics & Applied Nanoscience, Faculty of Physics, Bielefeld University, Universitätsstrasse 25, 33615 Bielefeld, Germany; Clinic of Thoracic and Cardiovascular Surgery, Heart and Diabetes Center NRW, University Hospital of the Ruhr-University Bochum, Medical School OWL (University of Bielefeld), Georgstrasse 11, D-32545 Bad Oeynhausen, Germany

**Keywords:** Desmin, Genetic Cardiomyopathy, Dilated Cardiomyopathy, Intermediate Filaments, Arrhythmia, Cytoskeleton

## Abstract

**Background:** Desmin is a muscle-specific intermediate filament protein crucial for maintaining cardiomyocyte structural integrity connecting multi-protein complexes and organelles. While *DES* mutations are known to cause various (cardio)myopathies, many rare variants remain classified as variants of uncertain significance (VUS).

**Methods:** We generated expression plasmids for 93 VUS located in the 1B domain and assessed filament formation in multiple cell lines, including cardiomyocytes derived from induced pluripotent stem cells. Filament assembly of purified wild-type and mutant desmin was analyzed using atomic force microscopy (AFM). Sequencing of 399 patients with severe dilated cardiomyopathy (DCM) identified the DES-p.L187P variant in one individual. Desmin localization in explanted myocardial tissue from this patient was examined via immunohistochemistry (IHC).

**Results:** Four variants (p.L159P, p.R163P, p.L187P, and p.E197del) caused filament formation defects, disrupting assembly even when co-expressed with wild-type desmin—consistent with dominant inheritance. AFM revealed that these mutations impaired filament formation, resulting in small desmin complexes, while wild-type desmin formed regular filaments. Systematic proline substitutions across the 1B domain showed that insertions at hydrophobic a- and d-sites disrupted filament assembly, whereas others had minimal impact. IHC confirmed desmin disorganization in myocardial tissue from a *DES-*p.L187P mutation carrier with DCM.

**Conclusion:** The atlas of cardiomyopathy-associated desmin mutations represents a significant step toward improving the clinical interpretation of *DES* variants associated with cardiomyopathies. Our data provide robust evidence, that four variants of previously unknown significance listed in the ClinVar database warrant reclassification as ‘likely pathogenic’ mutations based on their molecular effects. Specifically, we demonstrated that proline insertions – particularly at positions where hydrophobic amino acids contribute to the intermolecular interactions between alpha helices – lead to desmin filament assembly defects. These findings not only enhance our understanding of desmin-related cardiomyopathies but also offer a valuable resource for cardiologists and genetic counselors in guiding diagnosis, risk stratification and patient counseling.

## 1. Introduction

Beside environmental factors, cardiomyopathies can be caused by mutations in about 60 different genes including the *DES* gene, encoding the muscle-specific intermediate filament (IF) protein desmin ^1^. However, the classification and interpretation of rare cardiomyopathy-associated genetic variants is frequently limited by the lack of functional data ^2^. In consequence, the number of genetic variants of uncertain or unknown significance (VUS) is continuously increasing. Nevertheless, VUS leave frequently affected patients and their relatives dissatisfied and are consequently significant challenges for physicians and genetic counsellors involved in their health care.

Although it is known since the end of the 1990ies that pathogenic mutations in the *DES* gene cause different cardiomyopathies and combined or isolated skeletal myopathies ^3,4^, over 300 different VUS in the *DES* gene are currently listed in genetic disease databases like ClinVar (https://www.ncbi.nlm.nih.gov/clinvar/) and the ‘Human Gene Mutation Database’ (HGMD, https://www.hgmd.cf.ac.uk/ac/index.php). Desmin filaments connect several multi-protein complexes and cell organelles like the Z-bands, desmosomes, costameres, nuclei and mitochondria within the cardiomyocytes ^5,6^. Due to their structural flexibility ^7^, desmin filaments are highly important for the structural integrity of the cardiomyocytes ^8^.

Desmin consists of non-helical head and tail domains and a highly conserved rod domain, which is subdivided into 1A, 1B and coil-2 subdomains ^9^. The rod domains mediate a coiled-coil dimerization stabilized by a hydrophobic seam ^10^. Two of these dimers anneal into anti-parallel tetramers ^11^, which laterally assemble into unit-length filaments (ULFs) ^12^. Structural investigations using cryo-electron microscopy of the homologous vimentin filaments indicate the formation of five protofibrils and a central amyloid-like fiber mediated by head domain interactions ^13^.

Recently, we investigated the filament assembly of over 100 missense and small in-frame deletion variants localized in the desmin head and the 1A domain ^14,15^. These studies revealed a critical region for the filament assembly, spanning from the end of the head domain to the N-terminus of the 1A domain. Several pathogenic *DES* mutations in this region cause different cardiomyopathies by filament assembly _defects_ 14,16-20.

Due to the lack of functional data for the majority of VUS in the *DES* gene, we initiated this project to develop a comprehensive, detailed atlas of *DES* mutations leading to filament assembly defects. Here, we focus on the desmin 1B coil domain, in which about 90 different VUS are listed in genetic disease databases. We found that the majority of VUS within the 1B subdomain do not affect the desmin filament assembly. However, three amino acid exchanges against prolines (p.L159P, p.R163P and p.L187P) and one small in-frame deletion variant (p.E197del) disturbed the filament assembly in transfected cells including cardiomyocytes derived from induced pluripotent stem cells (iPSCs). These three proline variants formed aberrant cytoplasmic desmin aggregates and desmin-p.E197del formed mainly small fibrils. To verify these results, we expressed and purified these desmin mutants and investigated the filament assembly of recombinant desmin variants by atomic force microscopy (AFM). Since we observed that not all VUS inserting proline residues within the 1B subdomain disturb desmin filament formation, we conducted further investigations. Therefore, we systematically introduced proline residues at all positions in the 1B domain and analyzed the filament formation of them. Our findings revealed that proline residues within the hydrophobic heptad sequence have a detrimental effect.

In summary, we characterized the filament assembly of a set of 93 different VUS and 109 additional proline mutations (Σ=202 different variants) localized in the 1B domain of desmin. In future, these functional data could assist in the classification of additional *DES* variants and support genetic counselling of affected patients.

## 2. Material and Methods

All supporting data are included within the article and its supplemental material. The authors make their data, analytic methods and study material available to other researchers. The generated plasmids can be received from the corresponding author upon reasonable request. This study obtained institutional review board approval for the collection and IHC of explanted myocardial tissue and whole genome sequencing of the ethics review board (ERB) of the Ruhr-University Bochum (Bad Oeynhausen, Germany, 2024-1256 & AZ 2020-652 ). All patients gave informed consent. The study conforms with the declaration of Helsinki.

### 2.1 Analysis of Genetic Disease Databases

VUS in the *DES* gene affecting amino acids of the 1B desmin subdomain were received from the genetic disease databank ClinVar (https://www.ncbi.nlm.nih.gov/clinvar/) ^21^ and the Human Gene Mutation Database (hgmd.cf.ac.uk) ^22^ (November 2023).

### 2.2 Site-Directed Mutagenesis and Plasmid Generation

The generation of the plasmids pEYFP-N1-DES (Fig. S1), pmRuby-N1-DES (Fig. S2) and pET100D-TOPO-DES (Fig. S3) was previously described ^16,23^ . Most of the missense and small in-frame deletion variants were introduced into these plasmids using the QuikChange Lightning Site-Directed Mutagenesis Kit (Agilent Technologies,

Santa Clara, USA) or the Q5 Site-Directed Mutagenesis Kit (New England Biolabs, Ipswich, USA) in combination with specific oligonucleotides (Table S1, Microsynth, Balgach, Switzerland). The variants p.R189P, p.F211P, p.S231P and p.A252P were generated by overlap extension polymerase chain reaction (PCR) using Phusion High-Fidelity DNA polymerase (Thermo Fisher Scientific, , Waltham, USA) in combination with specific oligonucleotides (Table S1). PCR products were purified by agarose gel electrophoresis in combination with the GeneJET PCR Purification Kit (Thermo Fisher Scientific) and were afterwards cloned via *Xho*I and *Bam*HI into pEYFP-N1. The desmin encoding part of all plasmids were verified by Sanger sequencing (Macrogen, Amsterdam, Netherlands) (Fig. S4-S6).

### 2.3 Cell Culture

SW-13 and H9c2 cells were cultured at 37°C in Dulbecco’s Modified Eagle Medium (DMEM, Thermo Fisher Scientific) supplemented with 10% fetal calf serum (FCS, Thermo Fisher Scientific), penicillin and streptomycin. The cells were split two times per week using trypsin / ethylenediaminetetraacetic acid (EDTA, Thermo Fisher Scientific). Human iPSCs (NP00040-8, UKKi011-A, https://ebisc.org/UKKi011-A/), generated from a healthy male donor were cultured on vitronectin-coated cell culture flasks in Essential 8 medium (Thermo Fisher Scientific) as previously described ^24^.

### 2.4 Differentiation of Induced Pluripotent Stem Cells into Cardiomyocytes

Human iPSCs were differentiated into cardiomyocytes by modulating the Wnt-pathway using CHIR99021 and IWP2 as previously described in detail ^25^.

### 2.5 Cell Transfection

Cells were split one day before transfection and cultured in µ-Slide chambers (ibidi, Gräfelfing, Germany). Lipofectamin 3000 was used according to the manufacturer’s instructions for transfection using a plasmid – transfection reagent ratio of 1:3. The cells were cultured for 24 h after transfection under standard conditions (37°C, 5% CO_2_).

### 2.6 Fixation and Fluorescence Staining of Transfected Cells

The cells were washed with phosphate buffered saline (PBS). 4% Histofix (Carl Roth, Karlsruhe, Germany) was used for cell fixation (15 min, room temperature, RT). After washing with PBS, the cells were permeabilized using 0.1% Triton X-100 (15 min, RT) and were washed twice with PBS. Phalloidin conjugated to Texas Red (1:400, Thermo Fisher Scientific) and 4′,6-diamidino-2-phenylindole (DAPI, 1 µg/mL) were used for staining of F-actin or the nuclei. In iPSC-derived cardiomyocytes the Z-band protein α- actinin-2 was stained as a cardiomyocyte marker using a monoclonal mouse anti-α- actinin-2 antibody (1:100, Sigma Aldrich, #A7732) in combination with a secondary polyclonal goat anti-mouse immunoglobulin G antibody conjugated with Alexa Fluor 568 (1:100, Thermo Fisher Scientific, #A11004).

### 2.7 Confocal Microscopy Analysis

The TCS SP8 system (Leica Microsystems, Wetzlar, Germany) was used for confocal microscopy as previously described in detail ^15^. Three-dimensional cell stacks were generated and maximum intensity projections were generated with the Las X software (Leica Microsystems). The Huygens Essential software (Scientific Volume Imaging B.V., Hilversum, Netherlands) was used for deconvolution. For colocalization analysis of desmin-EYFP and -mRuby constructs, the Fiji software ^26^ was used in combination with the EzColocalization plugin ^27^.

### 2.8 Expression and Purification of Recombinant Desmin

Bacteria (*Escherichia coli*, One Shot BL21 (DE3) were transformed with pET100D- TOPO-DES expression plasmids according to the recommendations of the manufacturer (Thermo Fisher Scientific). After ampicillin selection (100 µg/mL), single colonies were picked and cultured in LB medium over night. These overnight cultures were used for inoculation. Recombinant desmin expression was induced by adding isopropyl β-D-1-thiogalactopyranoside (IPTG, 1 mM) when the OD600 reached a value about 0.4. The cultures were grown at 37°C for 4 h with intensive shaking. Afterwards, the bacteria were collected by centrifugation and were stored at -80°C.

Bacteria were thawed on ice and were incubated for 30 min in 30 mL lysis buffer (50 mM Tris-HCl, 25 mM sucrose, 1 mM EDTA, 10 mM dithiothreitol, 25 mM MgCl2, 0.1% Triton X-100, 1 mg/mL lysozym, 0.25 mg/mL DNase, pH 8.0) supplemented with proteinase inhibitor cocktail (Sigma-Aldrich, St. Louis, USA). After five washing steps, the pellet was solved in urea buffer (8 M urea, 20 mM Tris-HCl, pH 8.0). Ion exchange chromatography was performed using the ÄKTApurifier system (GE Healthcare, Chicago, USA) in combination with 5 mL HiTrap DEAE FF columns (Cytiva, Marlborough, USA). In a second purification step, immobilized metal affinity chromatography was performed using 5 mL HisTrap FF columns (Cytiva) as previously described ^15^. Recombinant desmin was stored at -80°C.

### 2.9 Desmin Preparation and Atomic Force Microscopy Analysis

Desmin filaments were prepared as previously described ^28^. Briefly, recombinant desmin was gradually dialyzed (1 h, RT) against DP-buffer (5 mM Tris-HCl, 1 mM DTT, pH 8.4) to decrease the urea concentration. The desmin concentration was then determined using absorption spectroscopy at 280 nm and adjusted to 0.3 g/l with DP-buffer. By adding an equal volume of assembly buffer (200 mM NaCl, 45 mM Tris-HCl, pH 7.0) filament assembly was initiated and the solution was incubated at 37°C for 1 h. Afterwards, 10 μL desmin solution were applied to freshly cleaved mica substrates and incubated for about 1 min. The samples were then carefully washed with water and dried under a gentle nitrogen flow. Topographic imaging under ambient conditions was performed in tapping mode using TAP300AI-G-Cantilevers (Budget Sensors, Sofia, Bulgaria) a JPK NanoWizard AFM (JPK Bruker, Berlin, Germany). Image analysis and processing was done with Gwyddion software (Version 2.62). Apart from first order plane correction all AFM scans are unprocessed.

### 2.10 Molecular Modelling and Protein Alignment

We used the molecular structure of the vimentin filaments ^13^ for homology modeling using the SWISS-MODEL server (https://swissmodel.expasy.org/) ^29^. PyMOL Molecular Graphics Version 2.52. (Schrödinger, New York, USA) was used for visualization of molecular desmin structure. Protein sequence alignments were generated by Clustal Omega (https://www.ebi.ac.uk/jdispatcher/msa/clustalo) ^30^ using the following desmin reference sequences: *Homo sapiens* (NP_001918.3), *Xenopus laevis* (NP_001080177.1), *Mus musculus* (NP_034173.1), *Rattus norvegicus* (NP_071976.2) and *Danio rerio* (NP_571038.2 and NP_001070920.1) and the human peripherin (NP_006253.2), vimentin (NP_003371.2) and GFAP (NP_002046.1) reference sequences.

### 2.11 Clinical Description of a DCM-Patient Carrying *DES*-p.L187P

A male patient in his 60’s with end-stage dilated cardiomyopathy (DCM, LVEF 18%), functional moderate mitral- and tricuspid regurgitation, secondary pulmonary hypertension, chronic renal insufficiency, and malignant ventricular arrhythmias had received a biventricular implantable cardioverter defibrillator (ICD). He was subsequently listed for heart transplantation (HTx). One months later the patient was transferred emergently from a rehabilitation facility after prolonged cardiopulmonary resuscitation. Persistent low-output syndrome prompted implantation of a Novacor left-ventricular assist device (LVAD) five days later. The early postoperative period was complicated by severe right-heart failure that necessitated inhaled nitric-oxide therapy.

Two months after LVAD implantation, the patient suffered a large left-hemispheric territorial infarction. Because of deep coma the patient was readmitted to the surgical intensive-care unit, where fulminant sepsis ensued. Hemorrhagic transformation of the infarct finally culminated in death. We performed a genetic analysis by next generation sequencing followed by Sanger sequencing and identified according to the ACMG guidelines ^2^ *DES-*p.L187P as a likely pathogenic variant.

### 2.12 Immunohistochemistry Analysis of Myocardial Tissue

Explanted left-ventricular myocardial tissue (stored at -80°C) from the *DES*-p.L187P mutation carrier and a piece from a rejected donor heart (control) was cut into 10 µm thick slices using a CM3050S cryomicrotome (Leica). 0.1% Triton X-100 (solved in PBS) was used for permeabilization (10 min at RT). 5% bovine serum albumin (solved in PBS) was used for 1 h at RT for blocking. Afterwards, the slices were washed two times with PBS and incubated with primary polyclonal goat anti-desmin antibodies over night at 4°C (AF3844, R&D Systems, Minneapolis, USA; 1:100). After several washing steps with PBS, the slices were incubated for 1 h at RT with secondary donkey anti-goat IgG conjugated with Alexa Fluor 555 (A21432, Thermo Fisher Scientific; 1:100) followed by several washing steps with PBS. Afterwards, the slices were incubated with phalloidin conjugated with Alexa Fluor 488 (A12379, Thermo Fisher Scientific; 1:400) for 40 min and DAPI (1 µg/mL) for 5 min at RT. Finally, the slices were washed several times with PBS. Dako Fluorescence Mounting Medium (Agilent) was used for embedding the slices. Afterwards, confocal microscopy in combination with deconvolution analysis was used for imaging using the TCS SP8 system (Leica).

### 2.13 Statistical Analysis

Four or more independent transfection experiments were performed per construct and per cell line and about 100 transfected cells were analyzed per transfection experiment. The non-parametric Kruskal-Wallis test followed by Dunn’s multiple comparison was used for statistical analysis using GraphPad Prism Version 10.2.3 (GraphPad Software, Boston, USA). All data are shown as means ± standard deviations (SD).

## Results

The 1B domains of type-3 IF proteins such as peripherin, GFAP, desmin and vimentin show significant homology (Fig. 1A). Furthermore, the desmin sequences across different vertebrate species are phylogenetically highly conserved underlining their functional relevance (Fig. 1B). The 1B domains mediate the intermolecular interaction within the tetrameric structure (Fig. 1C). In this study, we analyzed the ClinVar (https://www.ncbi.nlm.nih.gov/clinvar/, accessed April 17^th^, 2025) and the Human Gene Mutation Database (https://digitalinsights.qiagen.com/products-overview/clinical-insights-portfolio/human-gene-mutation-database/, accessed April 17^th^, 2025). From these databases, we identified 93 different VUS in the *DES* gene spread over the complete 1B domain (Fig. 1C). We included these VUS in our study if their minor allele frequency (MAF) was below 0.001 as reported by the Genome Aggregation Database (gnomAD v4.1.0, https://gnomad.broadinstitute.org/, accessed April 17^th^, 2025). We generated a set of 93 different expression plasmids encoding these VUS (Fig. S4). Next, we expressed both wild-type and mutant desmin, conjugated at the C-terminus with enhanced yellow fluorescent protein (EYFP) in SW- 13 cells. These cells lack endogenous desmin and other cytoplasmic IF proteins ^31^. In transiently transfected SW-13 cells, wild-type desmin-EYFP formed filaments with varying lengths and shapes (Fig. 1D). Likewise, wild-type desmin-EYFP assembled into filaments in H9c2 myoblasts and in cardiomyocytes derived from iPSCs (Fig. 1E-F). Both cell types express endogenous desmin.

**Figure 1.**
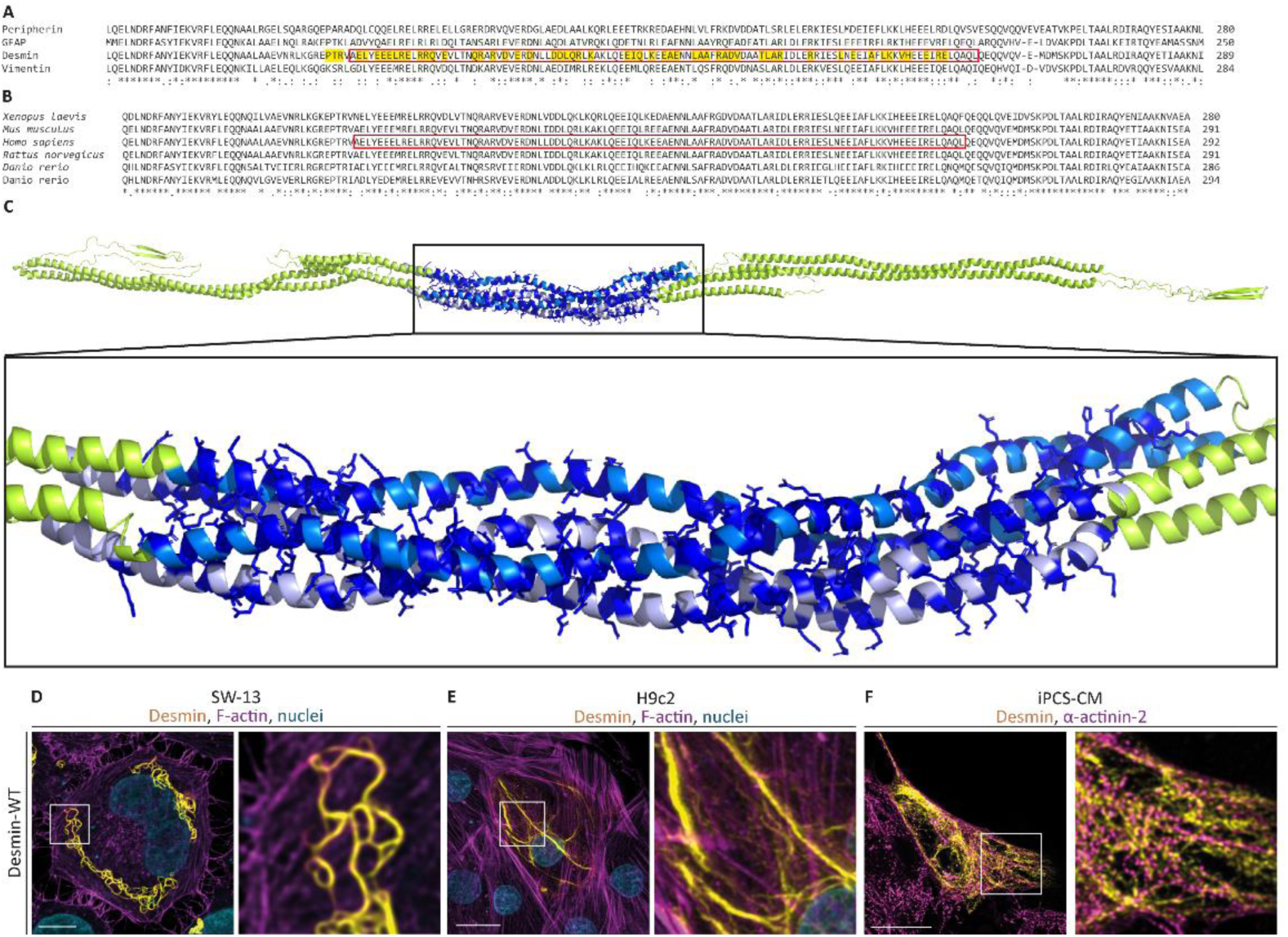
**(A)** Partial sequence alignment of different human type-3 intermediate filament proteins. The highly conserved 1A domain is shown by a red box and the positions of variants with unknown significance are highlighted in yellow. (**B**) Partial sequence alignment of desmin of different species. The highly conserved 1A domain is shown by a red box and the positions of variants with unknown significance are highlighted in yellow. (**A-B**) Identical amino acids are indicated with asterisks and similar ones are categorized with colons. (**C**) Structural overview of the desmin tetramer. The 1B domain is shown in light and marine blue. The positions of variants with unknown significance are highlighted in dark blue. Representative cell images of desmin wild-type transfected SW-13 (**D**), H9c2 cells (**E**) and iPSC-derived cardiomyocytes (**F**) are shown. Desmin is shown in yellow, F-actin or α-actinin in magenta, and the nuclei are shown in cyan. Scale bars represent 10 µm (**D**) or 20 µm (**E-F**).

89 of the desmin VUS in the 1B domain formed filamentous structures comparable to the wild-type desmin (Fig. 2A1-L6). However, three desmin missense mutants and one in-frame deletion mutation (p.L159P, p.R163P, p.L187P and p.E197del) disrupted the filament assembly leading to the formation of aberrant cytoplasmic desmin aggregates (Fig. 2-B6, -C2, -E8). In most transfected cells, desmin-p.E197del developed mixed filamentous and aggregate structures (Fig.2-F7). Notably, the AlphaMissense pathogenicity scores (AMS) ^32^ for desmin-p.L159P (0.990), -p.R163P (0.995) and - p.L187P (0.996) were the highest among the included desmin VUS within the 1B domain. However, other VUS with high AMS, such as p.K201N (0.978, Fig. 2-G3), p.T219I (0.988, Fig. 2-I3) or p.E246K (0.986, Fig. 2-K8) assembled into filamentous structures similar to those of wild-type desmin. Interestingly, ‘Rare Exome Variant Ensemble Learner’ (REVEL) scores fail to predict the aberrant desmin aggregation of specific VUS (Fig. 2) ^33^. For example, desmin-p.L159P and -p.E161G have similar REVEL scores (0.838 and 0.842), but desmin-p.L159P formed aggregates (Fig. 2-B6) and desmin-p.E161G assembled into filaments (Fig. 2-B8).

**Figure 2.**
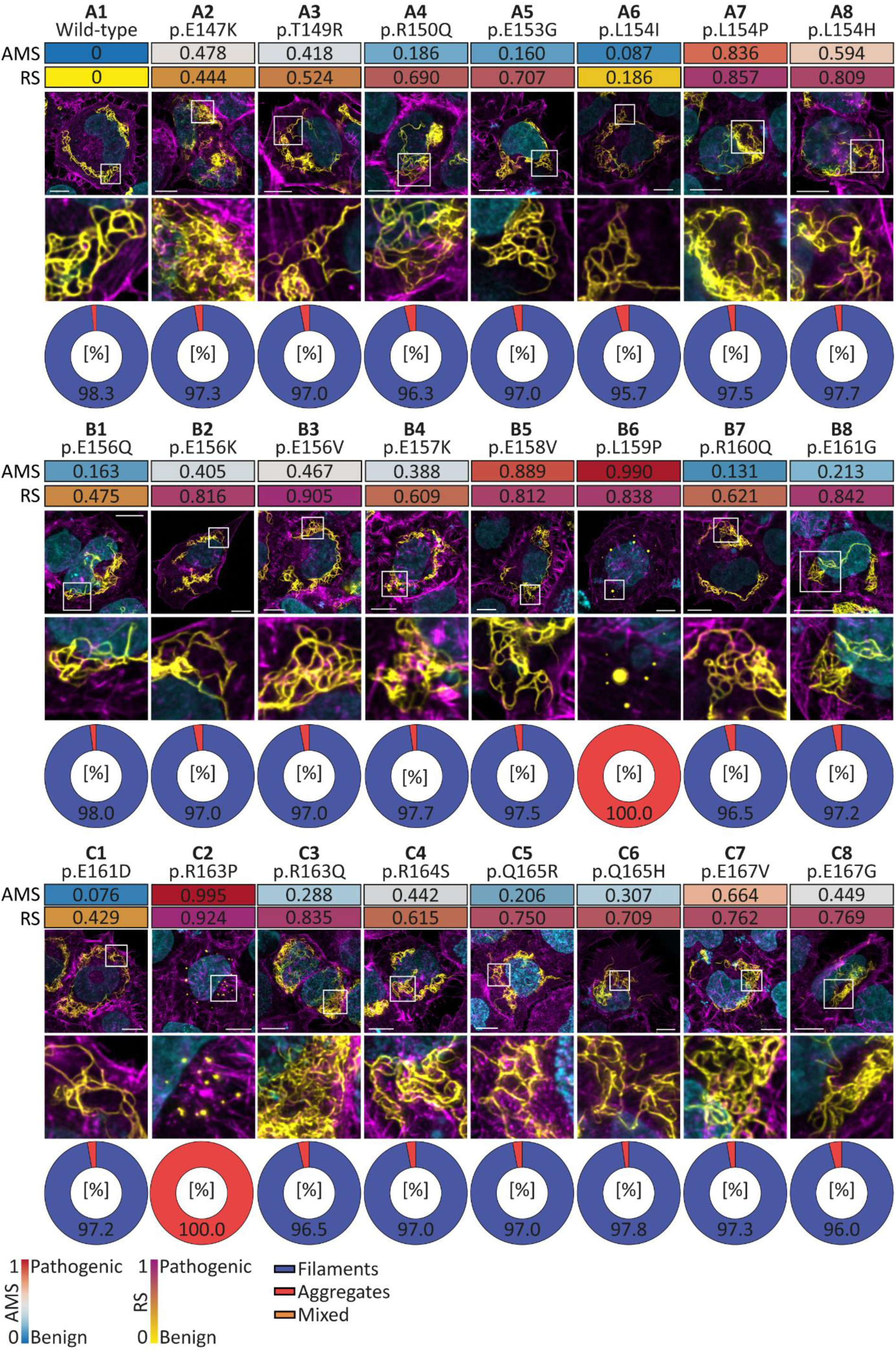
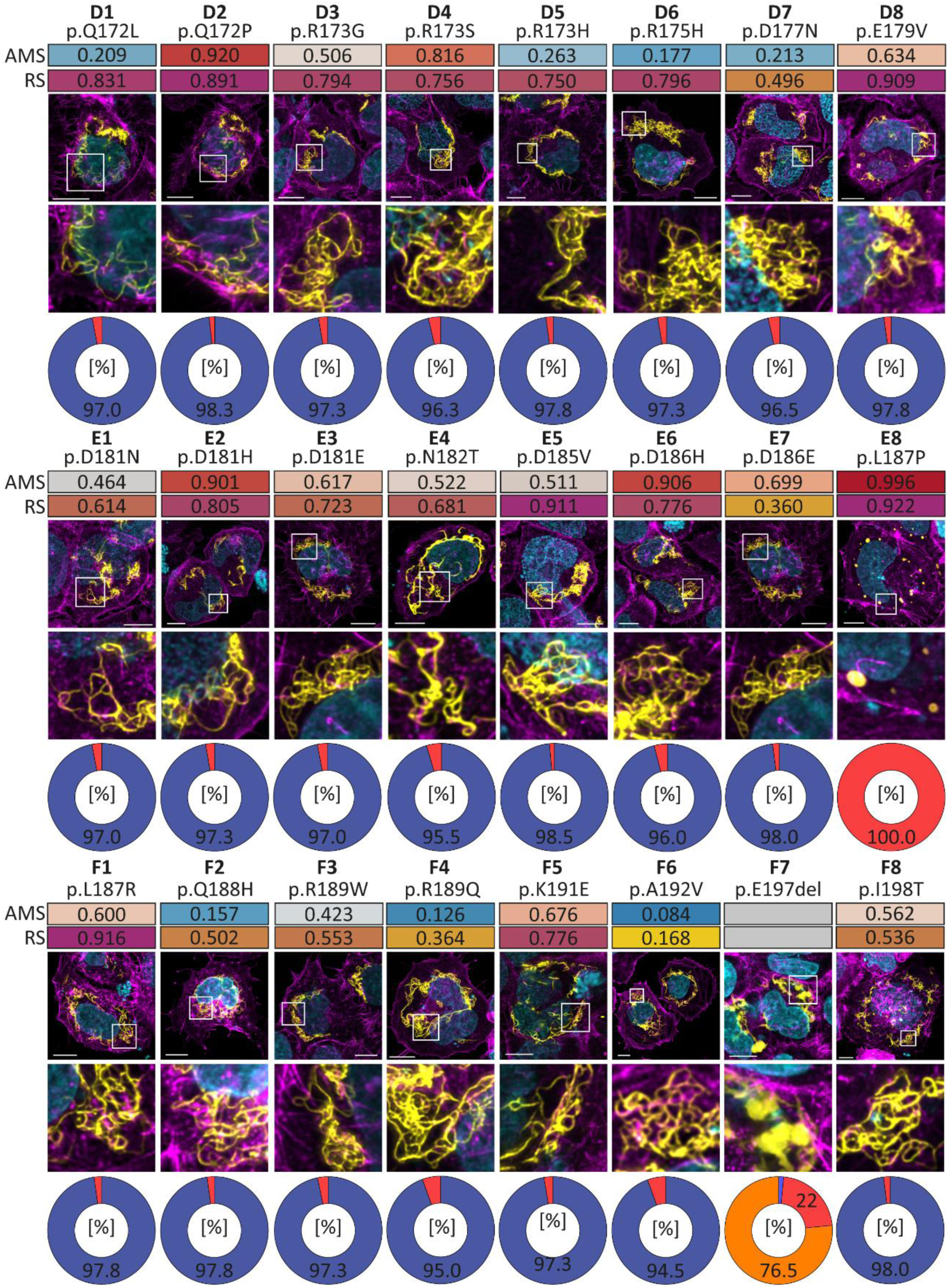
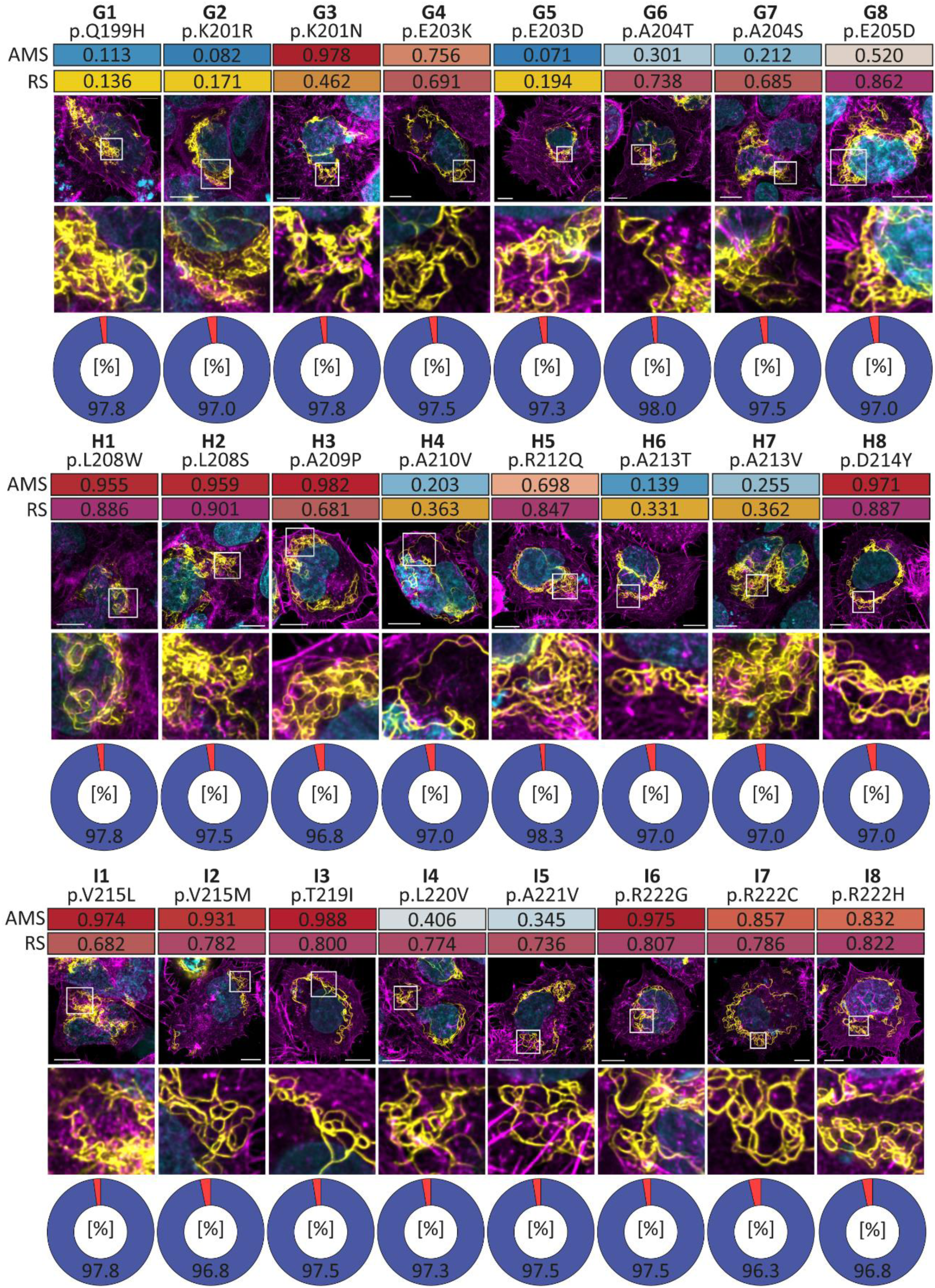
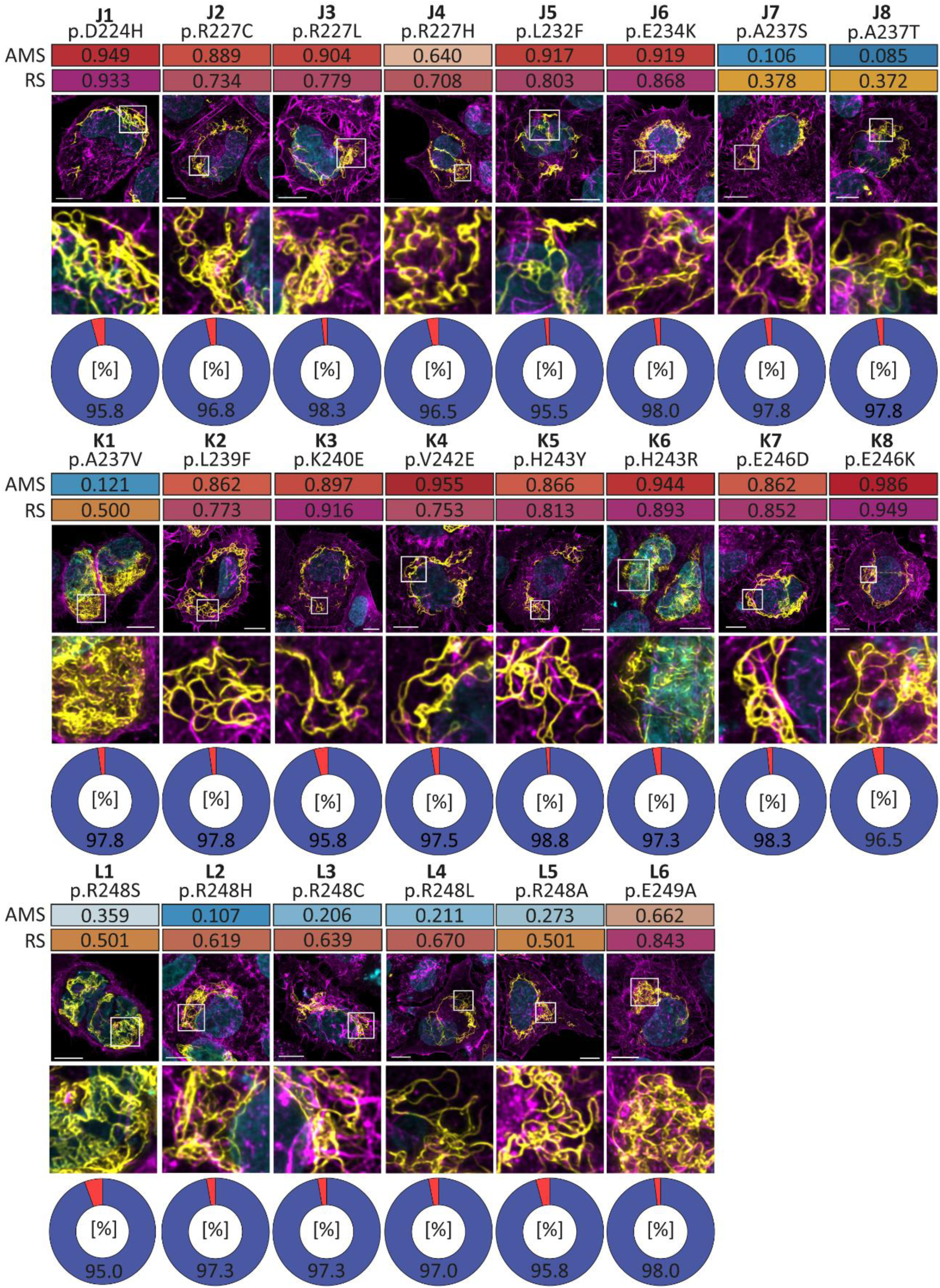
Representative cell images of wild-type **(A1)** and mutant **(A2-L6)** transfected SW-13 cells. Desmin-EYFP is shown in yellow, F-actin in magenta, and the nuclei are shown in cyan. Scale bars represent 10 µm. For each missense mutation the AlphaMissense Pathogenicity (AMS) ^32^ and the Rare Exome Variant Ensemble Learner (REVEL) ^33^ scores are indicated in heat maps ranging from 0-1. The percentages of the different cell phenotypes are summarized as pie charts (blue=filaments, red=aggregates or orange=mixed). Of note, the desmin mutants (p.L159P, **B6**; p.R163P, **C2**; p.L187P, **E8**; and p.E197del, **F7**) formed predominantly cytoplasmic aggregates, whereas most other VUS localized in the 1B domain formed similar filaments like the wild-type transfected SW-13 cells.

We expressed desmin-p.L159P, -p.R163P, -p.L187P and -p.E197del in H9c2 cells and in cardiomyocytes derived from iPSCs, since both cell types naturally express endogenous desmin (Fig. 3). In good agreement with our SW-13 transfection experiments, these three missense mutants (p.L159P, p.R163P and p.L187P) formed in H9c2 and iPSC-derived cardiomyocytes similar aberrant desmin aggregates (Fig. 3B-I), whereas the in-frame deletion mutant -p.E197del predominantly developed small fibrils in most transfected cells (Fig. 3E, 3J).

**Figure 3.**
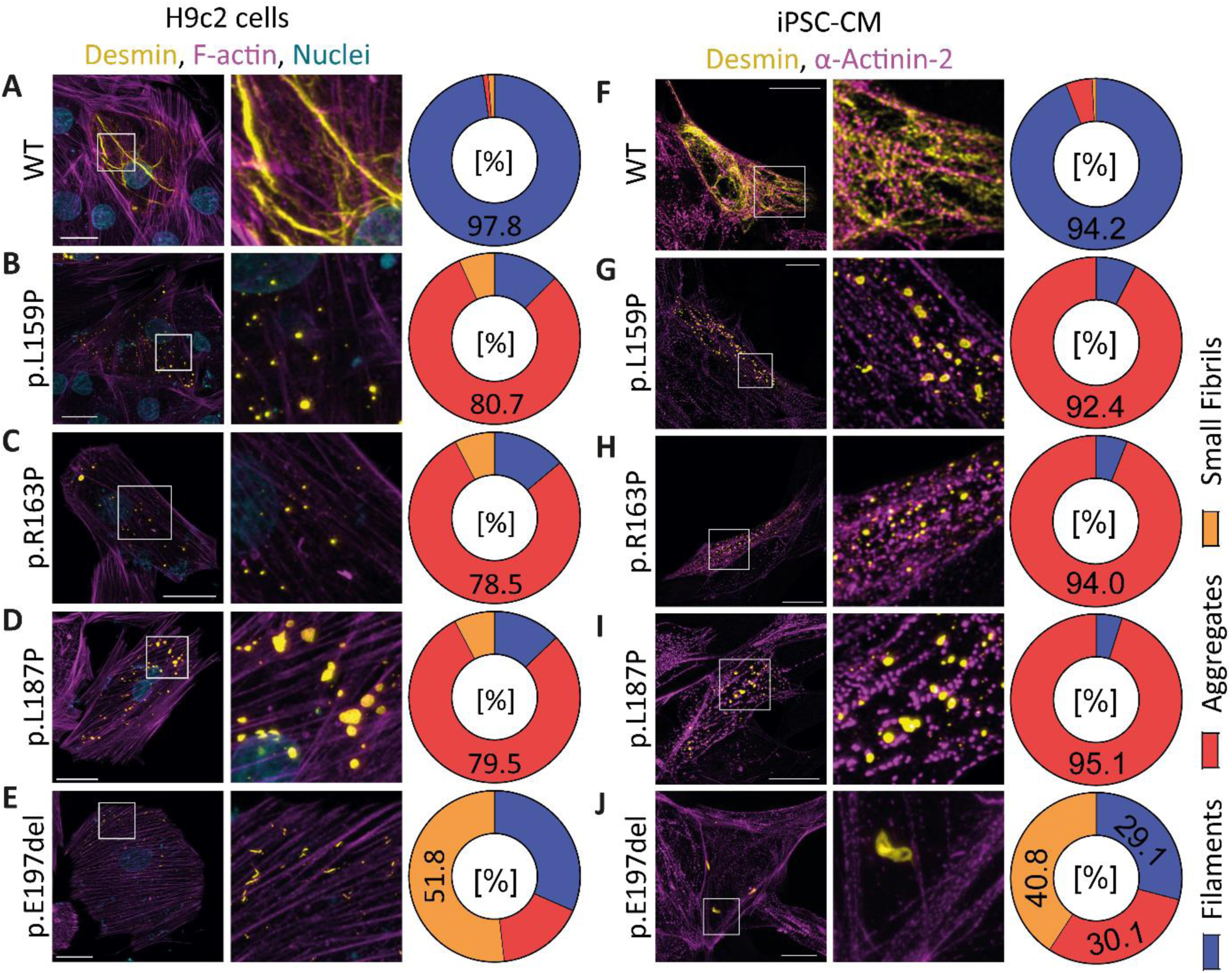
Representative cell images of H9c2 myoblasts **(A-E)** and iPSCs-derived cardiomyocytes (CM, **F-J**). Desmin-EYFP is shown in yellow, F-actin or α-Actinin-2 in magenta, and the nuclei are shown in cyan. Scale bars represent 20 µm. The percentages of the different cell phenotypes are summarized as pie charts (blue=filaments, red=aggregates or orange=small fibrils).

Following, recombinant wild-type and mutant desmin (p.L159P, p.R163P, p.L187P and p.E197del) were purified using ion exchange chromatography (IEC) in combination with immobilized metal affinity chromatography (IMAC). Filament assembly was analyzed by AFM revealing that wild-type desmin formed filaments with varying lengths (Fig. 4A). In contrast, the four desmin mutants failed to assemble into regular desmin filaments, producing only small structures (approx. 50-100 nm, Fig. 4B-E). Consequently, these experiments demonstrated that the four mutants (p.L159P, p.R163P, p.L187P and p.E197del) formed aberrant small molecular structures, indicating an intrinsic defect in desmin filament assembly.

**Figure 4.**
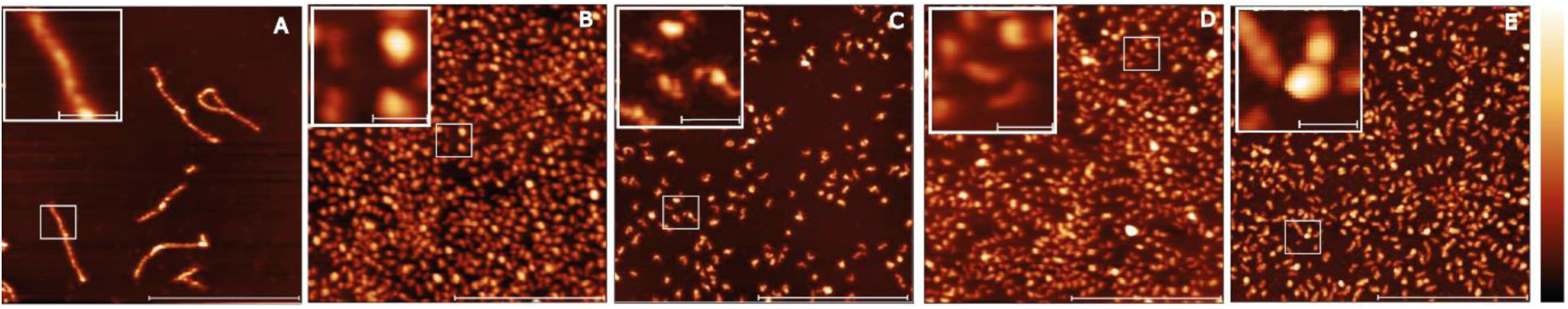
Representative atomic force microscopy topographic images of wildtype desmin and the mutants p.L159P, p.R163P, p.L187P and p.E197del. **(A)** Wildtype desmin assembles into long straight filaments with a typical length of about 600 nm. In contrast, the investigated desmin variants **(B)** p.L159P, **(C)** p.R163P, **(D)** p.L187P and **(E)** p.E197del form small aggregates and protofilaments with a size of 50-100 nm. The lateral scale bars correspond to 1 µm in the overview and 100 nm in the insets, while the color bar corresponds to a height of **(A)** 8 nm, **(B, E)** 18 nm, **(C)** 13 nm and **(D)** 15 nm.

Most desmin mutation carriers show a heterozygous genotype ^5^. Therefore, we performed co-transfection experiments of wild-type and mutant desmin to model the patient’s condition. In the control experiments, wild-type desmin fused either with EYFP or with mRuby formed typical filamentous networks, where both desmin forms are colocalized (Fig. 5A and S7). The proline missense mutations (p.L159P, p.R163P and p.L187P) formed, when co-expressed with wild-type desmin, predominantly aberrant cytoplasmic desmin aggregates consisting of both desmin forms (Fig. 5B-D and S7). In co-expression experiments, the deletion mutant desmin-p.E197del formed with wild-type desmin aggregates, filaments or mixtures of both (Fig. 5E and S7). However, these different structures contained mutant and wild-type desmin.

**Figure 5.**
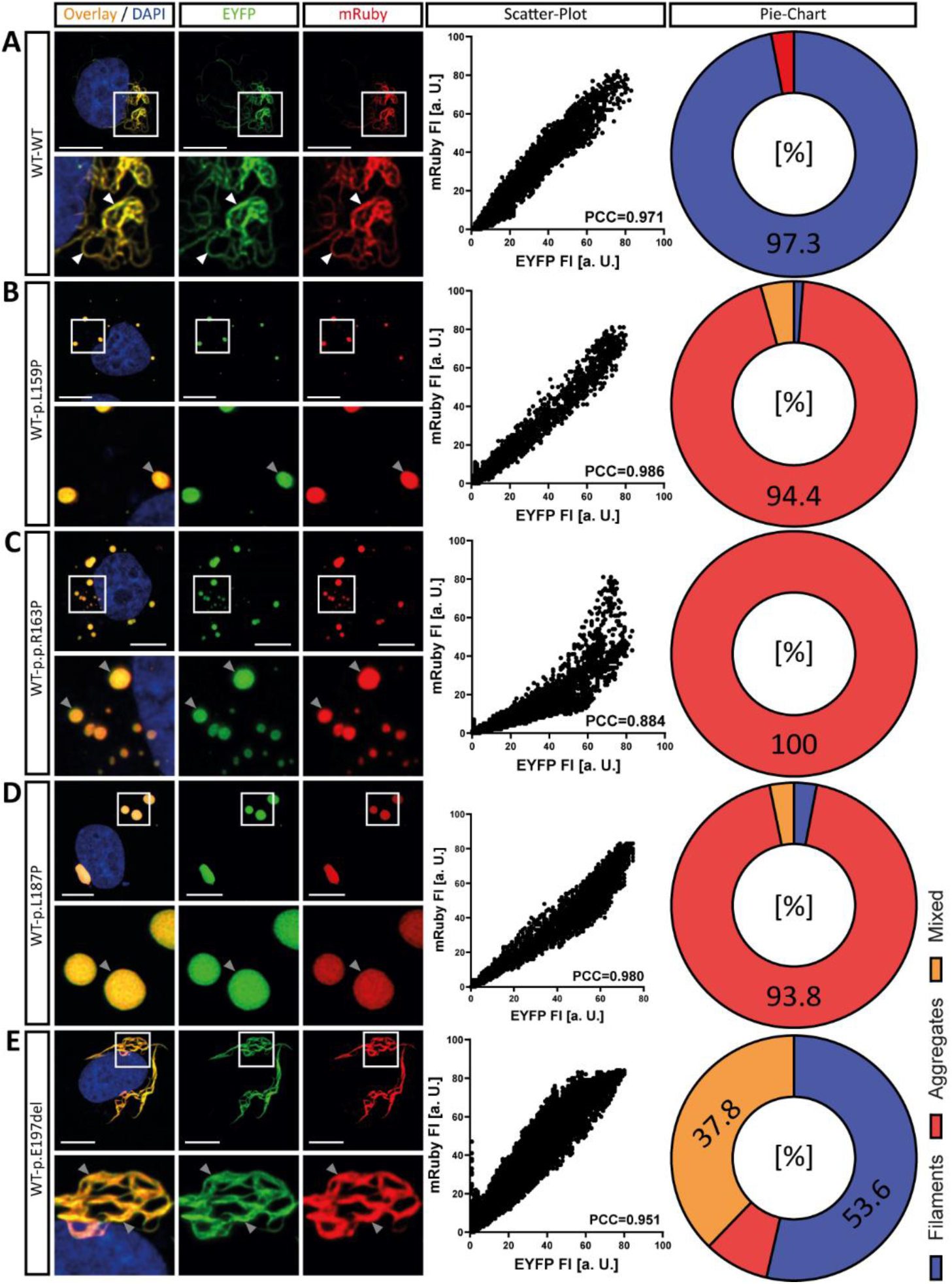
Double transfection experiments. Representative images of double-transfected SW-13 cells expressing wild-type desmin (mRuby-labeled, red) and either **(A)** wild-type or **(B-E)** mutant desmin (EYFP-labeled, green) are shown. Colocalization appears as a yellow overlay, with nuclei stained in blue. Scale bars: 10 µm. Scatter plots of mRuby and EYFP signal intensities were used to calculate the Pearson correlation coefficient (PCC). The distribution of cell phenotypes - desmin filaments, aggregates, or mixed structures - is summarized in pie charts.

Notably, our *DES* VUS screen revealed three proline variants (p.L154P, p.Q172P and p.A209P) that did not impair desmin filament assembly (Fig. 2-A7,-D2 and -H3), despite the detrimental effects observed with other proline substitutions (p.L159P, p.R163P and p.L187P). This raised the question whether the specific position of an inserted proline within the 1B domain plays a crucial role in inhibiting desmin filament assembly. To address this question, we systematically inserted proline residues at each position of the 1B domain (152-256) and analyzed their effects on filament formation in transfected SW-13 cells (Fig. 6). In contrast to expected potential effect that proline residues should impair filament formation due to their destabilizing influence on α-helices, we revealed that several proline mutations did not cause an aberrant desmin aggregation. However, the insertion of proline residues at the a- and d-positions, where hydrophobic amino acids stabilize the hydrophobic seam of wild-type desmin coiled-coil, frequently led to detrimental effects, resulting in aberrant desmin aggregation or the formation of abnormal thick filaments (Fig. 7). In contrast, proline residues at other positions within the heptad sequence do not disrupt the filament assembly in most cases (Fig. 7).

**Figure 6.**
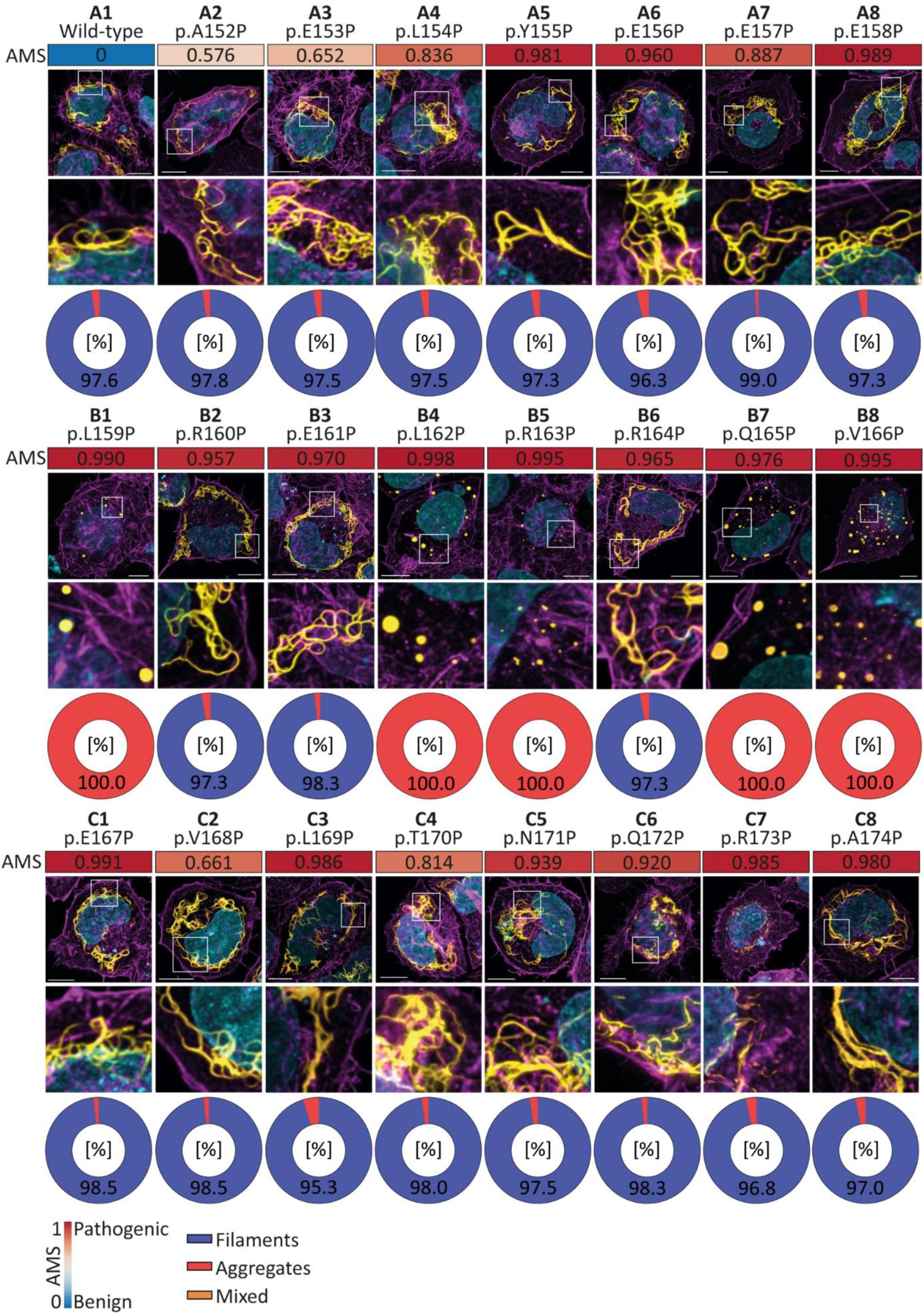
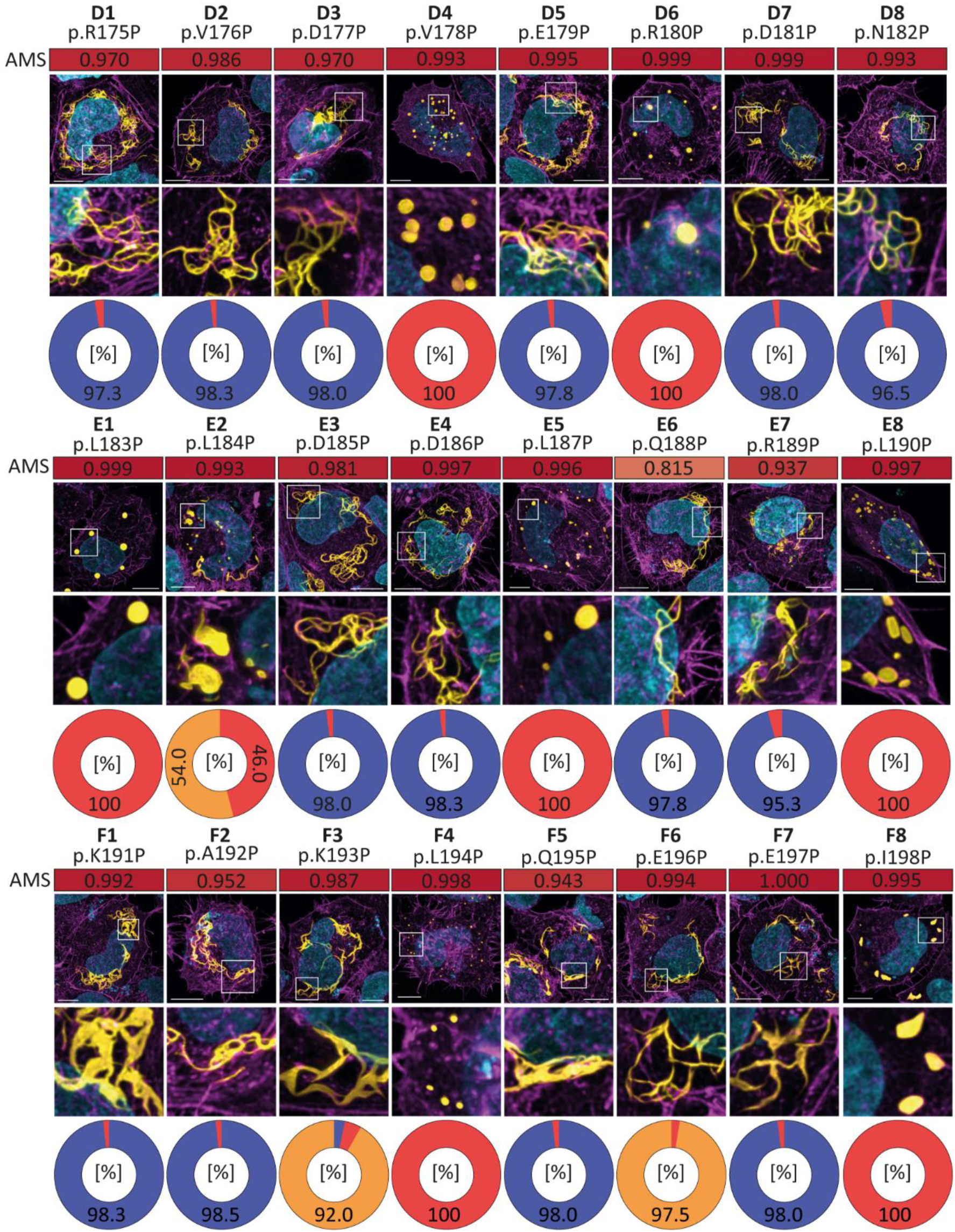
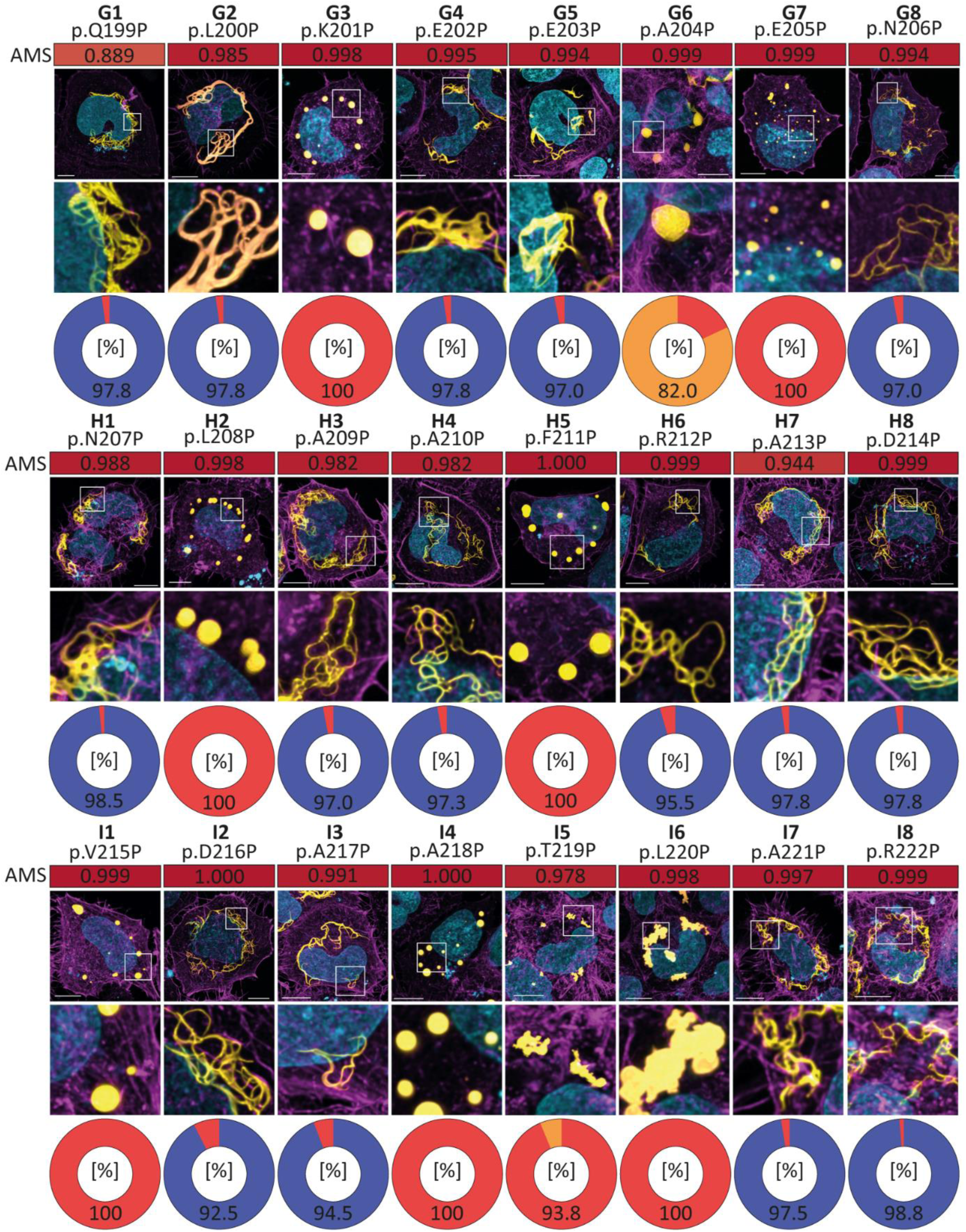
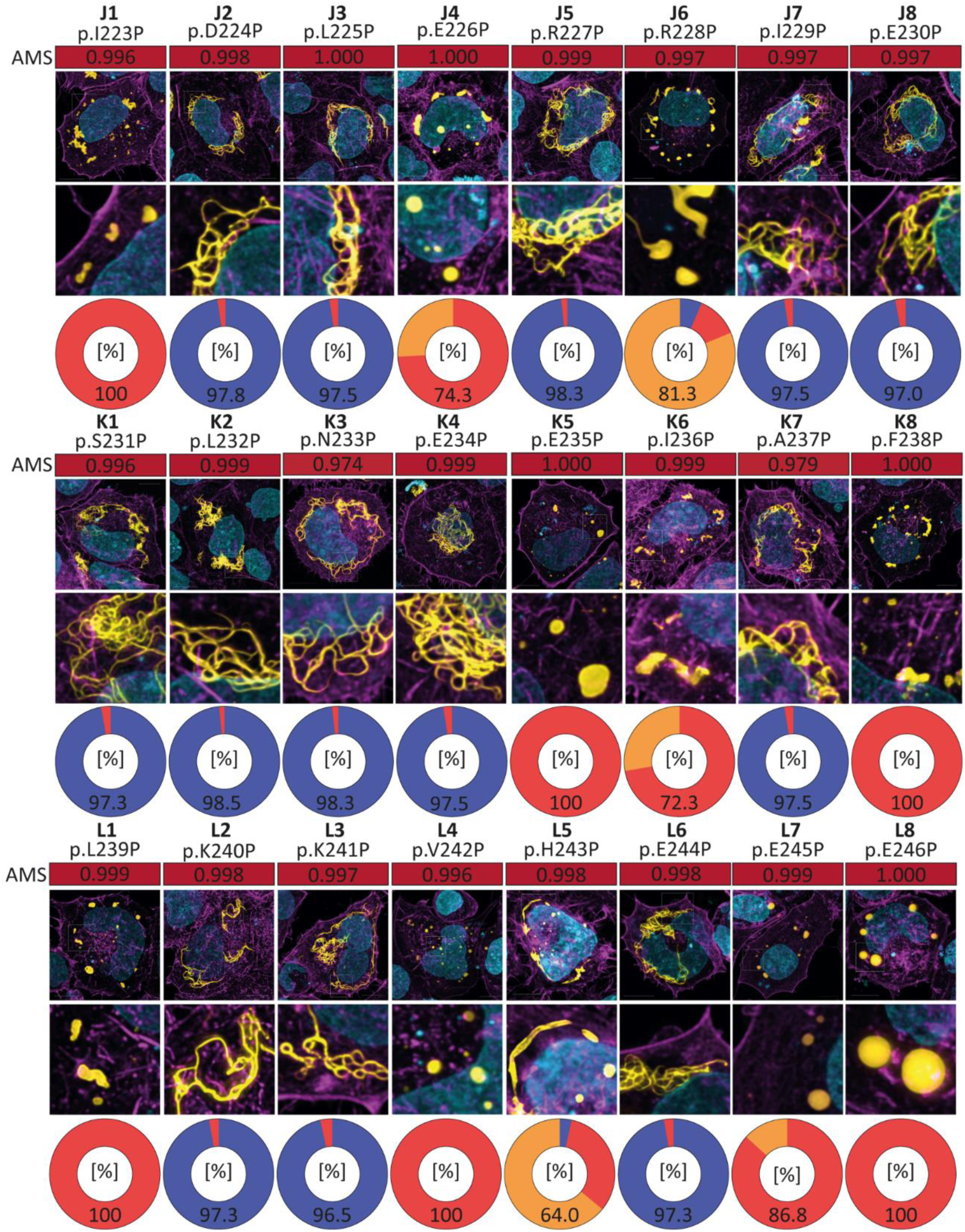
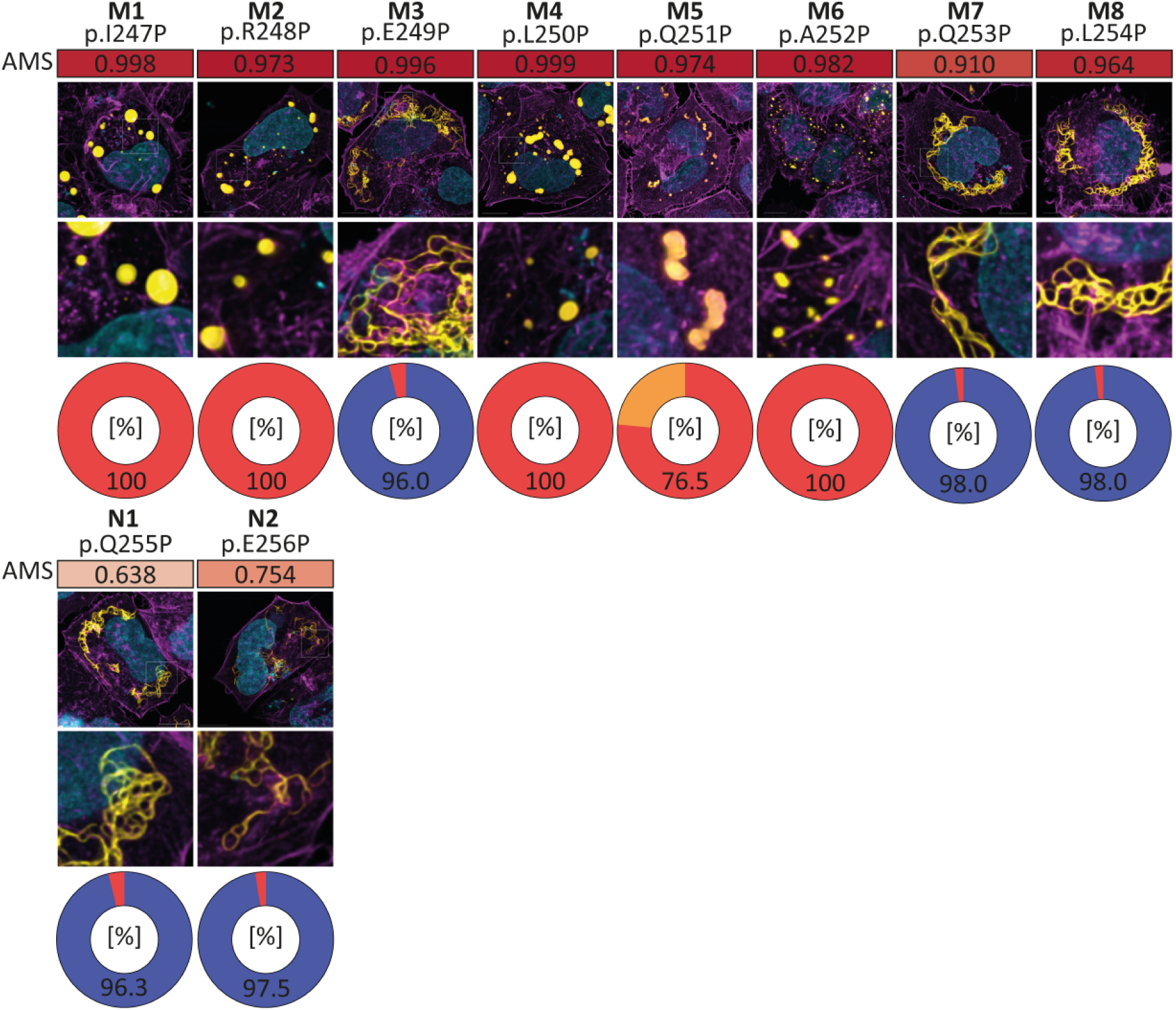
Proline screen of the desmin 1B domain. Representative cell images of wild-type **(A1)** and mutant **(A2-N2)** transfected SW-13 cells. Desmin-EYFP is shown in yellow, F-actin in magenta, and the nuclei are shown in cyan. Scale bars represent 10 µm. For each proline mutation the AlphaMissense Pathogenicity scores (AMS) ^32^ are indicated in heat maps ranging from 0-1. The percentages of the different cell phenotypes are summarized as pie charts (blue=filaments, red=aggregates or orange=aberrant thick filaments).

**Figure 7.**
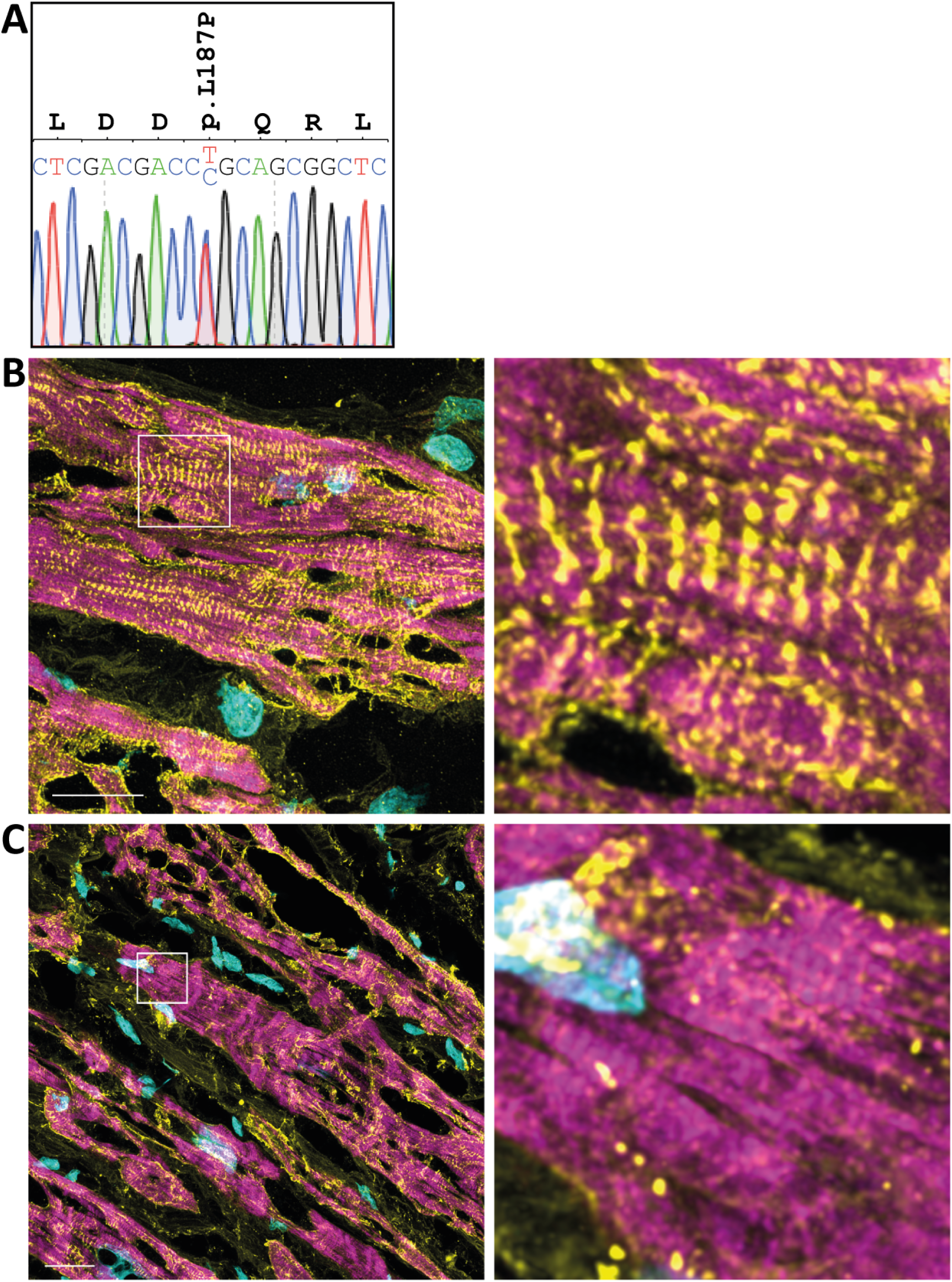
**(A)** Electropherogram of *DES*-p.L187P (c.560T>C) generated by Sanger sequencing using gDNA of the described DCM patient. Immunohistochemical analysis of left ventricular myocardial tissue (cyrosection, 10 µm) from the heterozygous *DES*- p.L187P patient **(B)** and a control sample (rejected donor heart) **(C)** were shown. All images are shown as maximum intensity projections. Desmin is shown in yellow, F- actin in magenta and the nuclei in cyan. Scale bars represent 20 nm.

Next, we analyzed whole genome data of 399 patients with end-stage DCM and medical need for HTx or LVAD implantation on *DES* variants. One patient carried the heterozygous variant *DES*-p.L187P (c.560T>C, Fig. 8A). However, this patient carried in addition a likely pathogenic *TTN* variant (p.Y33147*, c.99441C>A) and two rare VUS in the *LDB3* (p.D58N, c.172G>A) and *ALMS1* (p.R1818L, c.5453G>T) genes (Table S3). We used IHC in combination with confocal microscopy to investigate the structure of explanted myocardial tissue of this patient. Myocardial tissue from a rejected donor heart (non failing, NF) was used as a control (Fig. 8B). In contrast to the control, we found small aggregated and accumulated desmin and disorganized Z-bands and sarcomeres (Fig. 8C), all hallmarks of desminopathies, in case of the the *DES*-p.L187P mutation carrier. These data support our findings in cell culture data.

**Figure 8.**
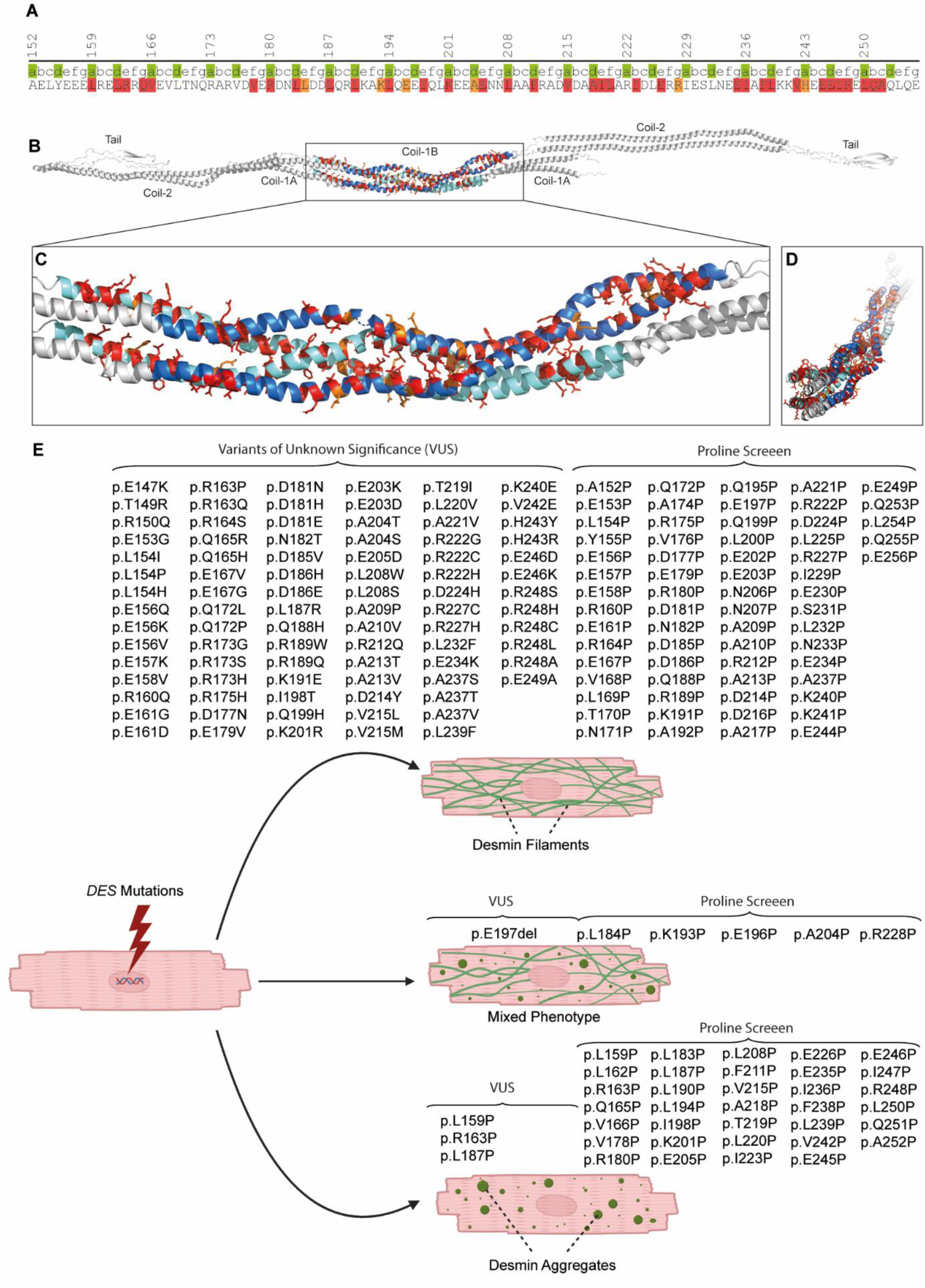
**(A)** Schematic overview about the proline screen. Sequence periodicity of the heptad (position a and d) is shown in green. The positions, where inserted proline residues disturbed the filament assembly are highlighted in red (aggregate formation) or orange (aberrant thick filaments). **(B-D)** Structural overview of the modelled desmin tetramer. The 1B desmin domains are shown in blue (forward dimer) or light blue (revers dimer). Head, 1A, coil-2 and tail domains are shown in grey. Amino acids, where exchanged proline mutations disturbed the filament assembly, are highlighted in red (aggregate formation) or orange (aberrant thick filaments). Of note, positions where proline residues impair the filament assembly are part of the hydrophobic heptad sequence. **(E)** Summarizing overview of aggregate or filament formation of the analyzed desmin mutations. Cardiomyocytes carrying expressing desmin variants (indicated by a flash) can show three different cellular phenotypes characterized by desmin filaments, a mixed morphology or aberrant cytoplasmic desmin aggregates. Cells with a mixed morphology show desmin filaments and aggregates. This figure was created with BioRender.com.

## Discussion

Accurate identification and classification of pathogenic genetic mutations associated with cardiomyopathies is essential for early therapeutic intervention and effective genetic counselling of affected patients and their relatives ^34^. Various bioinformatic in-silico tools, such as AlphaMissense, have been applied in cardiovascular genetics to predict the impact of specific genetic variants ^35^. Despite their predictive capabilities, these algorithms are currently limited by unknown accuracy of training data affecting their reliability ^35^. Accordingly, computational prediction tools are currently regarded only as supporting evidence within the guidelines of the American College of Medical Genetics and Genomics, whereas functional data can serve as a strong criterion for classification of genetic variants ^2^. However, the absence of functional data for most genetic variants currently hinders their interpretation. The more detailed framework provided by Brnich *et al.* refines the functional analysis of genetic variants ^36^. We followed their recommendations and provide a decision tree for the evaluation of functional data based on desmin filament assembly assays (Fig. S8).

Although mutations in the *DES* gene have been recognized as causes of various cardiomyopathies for over 20 years ^3,4^, predicting the pathogenic impact of specific rare VUS in the *DES* gene remains still challenging. Recently, we identified a hotspot for pathogenic *DES* mutations at the junction between the head and the 1A domain of desmin, leading to severe filament assembly defects ^14,15,37^. Desmin filaments play a crucial role in maintaining structural integrity of cardiomyocytes by connecting multi- protein complexes, such as the cardiac desmosomes, Z-bands and costameres, to various cellular organelles ^38^. In consequence, *DES* variants, that disrupt IF formation, are regarded as pathogenic, disease-causing mutations ^39,40^. Some pathogenic variants in the highly conserved 1B subdomain of desmin have been previously described ^41,42^. In this study, we systematically screened 93 distinct VUS within the desmin 1B domain for filament assembly defects, currently listed in the ClinVar or the HGMD databases (https://www.ncbi.nlm.nih.gov/clinvar/ and https://www.hgmd.cf.ac.uk/ac/index.php). Among these VUS, we identified three missense mutations (p.L159P, p.R163P and p.L187P) and one small in-frame deletion (p.E197del) leading to aberrant cytoplasmic desmin aggregates, which may contribute to reclassification of these *DES* variants. Notably, cytoplasmic IF proteins show a tissue specific expression pattern ^43^. Although desmin is the major cytoplasmic muscle specific IF protein, synemin and syncoilin are likewise expressed in cardiomyocytes ^38,44^. However, both IF proteins are unable to form regular IFs without the co-expression of desmin and cannot compensate the function of desmin. *Des* deficient mice develop severe cardiomyopathy underlining the unique function of desmin ^45,46^.

Of note, several pathogenic DCM-associated mutations have been reported at the homologous positions p.L85 and p.R89 (corresponding to p.L159 and p.R163 in desmin) of the homologous IF protein lamin A/C (*LMNA*) ^47,48^, suggesting that these sites are particularly sensitive to exchanges supporting their functional relevance for IF assembly.

In-frame deletions cause a twist of the helix explaining their detrimental impact as previously described ^49,50^. Proline residues are widely recognized as breakers of the α- helices, due to their lack of amide protons, which stabilize the protein backbone by h-bonds. Additionally, the bulkiness of the pyrrolidine ring causes steric hindrance and impairs the flexibility within α-helices ^51^. Interestingly, our VUS screen revealed that certain proline variants (p.L154P, p.Q172P and p.A209P) do not disrupt desmin filament formation and seem to be tolerated, maintaining filament integrity. This raises the question, at which position within the 1B domain proline residues have detrimental effects on IF assembly. Therefore, we performed a proline screen by inserting at each position of the 1B subdomain a proline residue. These experiments revealed that in most cases the proline residues located at an a- or d-position cause aberrant desmin aggregation, whereas most proline residues at other positions have minor or no effects on filament assembly (Fig. 7). In the desmin 1B subdomain, the a- and d-positions are predominantly composed of hydrophobic amino acids, such as leucine, isoleucine or valine, which mediate the intermolecular interactions between the α-helices within the desmin tetramer (Fig. 7).

In addition to our functional analysis, we analyzed whole genome data of 399 DCM patients for putative pathogenic or likely pathogenic variants in the *DES* gene. Of note, we identified in one patient with end-stage DCM the variant *DES*-p.L187P, which formed aberrant cytoplasmic desmin aggregates in our cell culture experiments. According to the ACMG guidelines ^2^, this variant fulfills two moderate (PS3 and PP3) and two supporting criteria of pathogenicity (PP2 and PM2), leading to its classification as a likely pathogenic variant (Table S3 and S4). However, this patient carried in addition a likely pathogenic nonsense mutation in the *TTN* gene, encoding the giant sarcomere protein titin. *TTN* nonsense mutations are known to cause frequently DCM ^52^. Therefore, a digenetic inheritance of both mutations in this DCM patient is likely. However, since the *DES* and *TTN* genes are both located on chromosome 2 and since DNA of the parents was not available, a maternal, paternal or mixed inheritance could not be excluded. Nevertheless, aberrant desmin aggregates and disorganized Z-bands, which are typical hallmarks for pathogenic *DES* mutations ^8^, were present in explanted myocardial tissue of this patient supporting our cell culture findings. The *in vitro* findings from filament assembly assays show a strong concordance with the pathogenic alterations identified in explanted myocardial tissue of a *DES*-p.L187P patient diagnosed with DCM.

In summary, we present here an atlas of cardiomyopathy associated desmin mutations within the 1B domain, which has the potential to enhance the clinical interpretation of rare *DES* variants in genetic diagnosis of non-ischemic cardiomyopathies. Moreover, it may serve as a valuable resource for genetic counseling of affected cardiomyopathy patients and their relatives.

## Limitations

Desminopathies are known for their broad clinical phenotypic spectrum affecting skeletal and cardiac muscle ^50,53,54^. Therefore, the presented *in vitro* data may not fully capture all aspects of desminopathies but focus on the myocardial impact caused by desmin filament assembly defects.

## Author Contributions

Conceptualization (AB); Data curation (SV, FK, and AB); Formal analysis (SV, FK, VW and AB); Funding acquisition (AB and HM); Investigation (SV, FK, JR, MS, SH and AB); Project administration (AB); Clinical Investigations (MAD, JG); Resources (JG, DA); Supervision (AB and VW); Visualization (SV, FK, VW and AB); Roles/Writing - original draft (AB); and Writing - review & editing (all authors).

## Supplemental Materials

Tables S1-S4.

Figure S1-S8.

## Non-standard Abbreviations and Acronyms

AFM: Atomic Force Microscopy
DAPI: 4′,6-Diamidino-2-phenylindole
DMEM: Dulbecco’s Modified Eagle Medium
EDTA: Ethylenediaminetetraacetic Acid
EYFP: Enhanced Yellow Fluorescent Protein
FCS: Fetal Calf Serum
HTx: Heart Transplantation
IF: Intermediate Filaments
ICD: Implantable Cardioverter Defibrillator
iPSCs: Induced Pluripotent Stem Cells
PBS: Phosphate Buffered Saline
PCR: Polymerase Chain Reaction
LVEF: Left-Ventricular Ejection Fraction
LVAD: Left-Ventricular Assist Device
MAF: Minor Allele Frequency
SD: Standard Deviation
ULFs: Unit Length Filaments
VUS: Variant of Uncertain Significance

## Data Availability

All supporting data are included within the article and its supplemental material. The authors make their data, analytic methods and study material available to other researchers. The generated plasmids can be received from the corresponding author upon reasonable request.

