## Supplemental Material for "Atlas of cardiomyopathy associated *DES* (desmin) mutations: Functional insights into the critical 1B domain"

**Table S1.** Overview about the used oligonucleotides.

| Name | Sequence 5‘-3‘ | Application |
| --- | --- | --- |
| CMV_for | CGCAAATGGGCGGTAGGCGTG | Sanger Sequencing |
| EGFP_N_rev | GCTTGCCGTAGGTGGCATC | Sanger Sequencing |
| T7_for | TAATACGACTCACTATAGGG | Sanger Sequencing |
| T7_rev | GCTAGTTATTGCTCAGCGGT | Sanger Sequencing |
| DES_Overlap_for | CGGACTCAGATCTCGAGGCCGTC | Overlap-PCR |
| DES_Overlap_rev | GTCAGCTTGCCGTAGGTGGCATC | Overlap-PCR |
| DES_E147K_for | CTCAAGGGCCGC**A**AGCCGACGCGA G | SDM (QCL) |
| DES_E147K_rev | CTCGCGTCGGCT**T**GCGGCCCTTGA G | SDM (QCL) |
| DES_T149R_for | GCCGCGAGCCGA**G**GCGAGTGG C | SDM (QCL) |
| DES_T149R_rev | GCCACTCGC**C**TCGGCTCGCGG C | SDM (QCL) |
| DES_R150Q_for | CGAGCCGACGC**A**AGTGGCCGAGC | SDM (QCL) |
| DES_ R150Q _rev | GCTCGGCCACT**T**GCGTCGGCTCG | SDM (QCL) |
| DES_ E153G _for | CGCGAGTGGCCG**G**GCTCTACGAGGA | SDM (QCL) |
| DES_ E153G _rev | TCCTCGTAGAGC**C**CGGCCACTCGCG | SDM (QCL) |
| DES_L154I_for | GCGAGTGGCCGAG**A**TCTACGAGGAGGA | SDM (QCL) |
| DES_L154I_rev | TCCTCCTCGTAGA**T**CTCGGCCACTCGC | SDM (QCL) |
| DES_L154P_for | CGAGTGGCCGAGC**C**CTACGAGGAGGAG | SDM (QCL) |
| DES_L154P_rev | CTCCTCCTCGTAG**G**GCTCGGCCACTCG | SDM (QCL) |
| DES_L154H_for | CGAGTGGCCGAGC**A**CTACGAGGAGGAG | SDM (QCL) |
| DES_L154H_rev | CTCCTCCTCGTAG**T**GCTCGGCCACTCG | SDM (QCL) |
| DES_E156Q_for | GCCGAGCTCTAC**C**AGGAGGAGCTGC | SDM (QCL) |
| DES_E156Q_rev | GCAGCTCCTCCT**G**GTAGAGCTCGGC | SDM (QCL) |
| DES_E156K_for | GGCCGAGCTCTAC**A**AGGAGGAGCTGCG | SDM (QCL) |
| DES_E156K_rev | CGCAGCTCCTCCT**T**GTAGAGCTCGGCC | SDM (QCL) |
| DES_E156V_for | CCGAGCTCTACG**T**GGAGGAGCTGCG | SDM (QCL) |
| DES_E156V_rev | CGCAGCTCCTCC**A**CGTAGAGCTCGG | SDM (QCL) |
| DES_E157K_for | CGAGCTCTACGAG**A**AGGAGCTGCGGGA | SDM (QCL) |
| DES_E157K_rev | TCCCGCAGCTCCT**T**CTCGTAGAGCTCG | SDM (QCL) |
| DES_E158V_for | CTCTACGAGGAGG**T**GCTGCGGGAGCTG | SDM (QCL) |
| DES_E158V_rev | CAGCTCCCGCAGC**A**CCTCCTCGTAGAG | SDM (QCL) |
| DES_L159P_for | CGAGGAGGAGC**C**GCGGGAGCTGC | SDM (QCL) |
| DES_L159P_rev | GCAGCTCCCGC**G**GCTCCTCCTCG | SDM (QCL) |
| DES_R160Q_for | GGAGGAGCTGC**A**GGAGCTGCGGC | SDM (QCL) |
| DES_R160Q_rev | GCCGCAGCTCC**T**GCAGCTCCTCC | SDM (QCL) |
| DES_E161G_for | GGAGCTGCGGG**G**GCTGCGGCGCC | SDM (QCL) |
| DES_E161G_rev | GGCGCCGCAGC**C**CCCGCAGCTCC | SDM (QCL) |
| DES_E161D_for | GAGCTGCGGGA**T**CTGCGGCGCCA | SDM (QCL) |
| DES_E161D_rev | TGGCGCCGCAG**A**TCCCGCAGCTC | SDM (QCL) |
| DES_R163P_for | GCGGGAGCTGC**C**GCGCCAGGTGG | SDM (QCL) |
| DES_R163P_rev | CCACCTGGCGC**G**GCAGCTCCCGC | SDM (QCL) |
| DES_R163Q_for | GCGGGAGCTGC**A**GCGCCAGGTGG | SDM (QCL) |
| DES_R163Q_rev | CCACCTGGCGC**T**GCAGCTCCCGC | SDM (QCL) |
| DES_R164S_for | GGGAGCTGCGG**A**GCCAGGTGGAG | SDM (QCL) |
| DES_R164S_rev | CTCCACCTGGC**T**CCGCAGCTCCC | SDM (QCL) |
| DES_Q165R_for | GCTGCGGCGCC**G**GGTGGAGGTGC | SDM (QCL) |
| DES_Q165R_rev | GCACCTCCACC**C**GGCGCCGCAGC | SDM (QCL) |
| DES_Q165H_for | GCTGCGGCGCCA**T**GTGGAGGTGCTC | SDM (QCL) |
| DES_Q165H_rev | GAGCACCTCCAC**A**TGGCGCCGCAGC | SDM (QCL) |
| DES_E167V_for | GTTAGTGAGCACC**A**CCACCTGGCGCCG | SDM (QCL) |
| DES_E167V_rev | CGGCGCCAGGTGG**T**GGTGCTCACTAAC | SDM (QCL) |
| DES_E167G_for | CGGCGCCAGGTGG**G**GGTGCTCACTAAC | SDM (QCL) |
| DES_E167G_rev | GTTAGTGAGCACC**C**CCACCTGGCGCCG | SDM (QCL) |
| DES_Q172L_for | GCTCACTAACC**T**GCGCGCGCGCG | SDM (QCL) |
| DES_Q172L_rev | CGCGCGCGCGC**A**GGTTAGTGAGC | SDM (QCL) |
| DES_Q172P_for | GCTCACTAACC**C**GCGCGCGCGCG | SDM (QCL) |
| DES_Q172P_rev | CGCGCGCGCGC**G**GGTTAGTGAGC | SDM (QCL) |
| DES_R173G_for | CTCACTAACCAG**G**GCGCGCGCGTCG | SDM (QCL) |
| DES_R173G_rev | CGACGCGCGCGC**C**CTGGTTAGTGAG | SDM (QCL) |
| DES_R173S_for | CTCACTAACCAG**A**GCGCGCGCGTCG | SDM (QCL) |
| DES_R173S_rev | CGACGCGCGCGC**T**CTGGTTAGTGAG | SDM (QCL) |
| DES_R173H_for | CTCACTAACCAGC**A**CGCGCGCGTCGAC | SDM (QCL) |
| DES_R173H_rev | GTCGACGCGCGCG**T**GCTGGTTAGTGAG | SDM (QCL) |
| DES_R175H_for | CCAGCGCGCGC**A**CGTCGACGTCG | SDM (QCL) |
| DES_R175H_rev | CGACGTCGACG**T**GCGCGCGCTGG | SDM (QCL) |
| DES_D177N_for | GCGCGCGCGTC**A**ACGTCGAGCGC | SDM (QCL) |
| DES_D177N_rev | GCGCTCGACGT**T**GACGCGCGCGC | SDM (QCL) |
| DES_E179V_rev | AGGTTGTCGCGC**A**CGACGTCGACGC | SDM (QCL) |
| DES_D181N_for | CGACGTCGAGCGC**A**ACAACCTGCTCGA | SDM (QCL) |
| DES_D181N_rev | TCGAGCAGGTTGT**T**GCGCTCGACGTCG | SDM (QCL) |
| DES_D181H_for | GACGTCGAGCGC**C**ACAACCTGCTCG | SDM (QCL) |
| DES_D181H_rev | CGAGCAGGTTGT**G**GCGCTCGACGTC | SDM (QCL) |
| DES_D181E_for | CGTCGAGCGCGA**G**AACCTGCTCGAC | SDM (QCL) |
| DES_D181E_rev | GTCGAGCAGGTT**C**TCGCGCTCGACG | SDM (QCL) |
| DES_N182T_for | CGACGTCGAGCGCGACA**C**CCTGCTCGA | SDM (QCL) |
| DES_N182T_rev | TCGAGCAGG**G**TGTCGCGCTCGACGTCG | SDM (QCL) |
| DES_D185V_for | GACAACCTGCTCG**T**CGACCTGCAGCGG | SDM (QCL) |
| DES_D185V_rev | CCGCTGCAGGTCG**A**CGAGCAGGTTGTC | SDM (QCL) |
| DES_D186H_for | AACCTGCTCGAC**C**ACCTGCAGCGGC | SDM (QCL) |
| DES_D186H_rev | GCCGCTGCAGGT**G**GTCGAGCAGGTT | SDM (QCL) |
| DES_D186E_for | CCTGCTCGACGA**G**CTGCAGCGGCTC | SDM (QCL) |
| DES_D186E_rev | GAGCCGCTGCAG**C**TCGTCGAGCAGG | SDM (QCL) |
| DES_L187P_for | CTGCTCGACGACC**C**GCAGCGGCTCAAG | SDM (QCL) |
| DES_L187P_rev | CTTGAGCCGCTGC**G**GGTCGTCGAGCAG | SDM (QCL) |
| DES_L187R_for | CTGCTCGACGACC**G**GCAGCGGCTCAAG | SDM (QCL) |
| DES_L187R_rev | CTTGAGCCGCTGC**C**GGTCGTCGAGCAG | SDM (QCL) |
| DES_Q188H_for | CTGCTCGACGACCTGCA**T**CGGCTCAAGG | SDM (QCL) |
| DES_Q188H_rev | CCTTGAGCCG**A**TGCAGGTCGTCGAGCAG | SDM (QCL) |
| DES_R189W_for | CGACGACCTGCAG**T**GGCTCAAGGCCAA | SDM (QCL) |
| DES_R189W_rev | TTGGCCTTGAGCC**A**CTGCAGGTCGTCG | SDM (QCL) |
| DES_R189Q_for | GACGACCTGCAGC**A**GCTCAAGGC | Overlap-PCR |
| DES_R189Q_rev | GCCTTGAGC**T**GCTGCAGGTCGTC | Overlap-PCR |
| DES_K191E_for | CTGCAGCGGCTC**G**AGGCCAAGCTGC | SDM (QCL) |
| DES_K191E_rev | GCAGCTTGGCCT**C**GAGCCGCTGCAG | SDM (QCL) |
| DES_A192V_for | CAGCGGCTCAAGG**T**CAAGCTGCAGGAG | SDM (QCL) |
| DES_A192V_rev | CTCCTGCAGCTTG**A**CCTTGAGCCGCTG | SDM (QCL) |
| DES_E197del_for | TCTTCCTTCAACTGAATCTCCTGCAGCTTGGCC | SDM (QCL) |
| DES_E197del_rev | GGCCAAGCTGCAGGAGATTCAGTTGAAGGAAGA | SDM (QCL) |
| DES_I198T_for | GCCAAGCTGCAGGAGGAGA**C**TCAGTTGAAGGAAG | SDM (QCL) |
| DES_I198T_rev | CTTCCTTCAACTG**A**GTCTCCTCCTGCAGCTTGGC | SDM (QCL) |
| DES_Q199H_for | CTGCAGGAGGAGATTCA**T**TTGAAGGAAGAAGCAGA | SDM (QCL) |
| DES_Q199H_rev | TCTGCTTCTTCCTTCAA**A**TGAATCTCCTCCTGCAG | SDM (QCL) |
| DES_K201R_for | CAGGAGGAGATTCAGTTGA**G**GGAAGAAGCAGAGAAC | SDM (QCL) |
| DES_K201R_rev | GTTCTCTGCTTCTTCC**C**TCAACTGAATCTCCTCCTG | SDM (QCL) |
| DES_K201N_for | CAGGAGGAGATTCAGTTGAA**T**GAAGAAGCAGAGAACAATTT | SDM (QCL) |
| DES_K201N_rev | AAATTGTTCTCTGCTTCTTC**A**TTCAACTGAATCTCCTCCTG | SDM (QCL) |
| DES_E203K_for | CAGGAGGAGATTCAGTTGAAGGAA**A**AAGCAGAGAACAATTT | SDM (QCL) |
| DES_E203K_rev | AAATTGTTCTCTGCTT**T**TTCCTTCAACTGAATCTCCTCCTG | SDM (QCL) |
| DES_E203D_for | AGGAGGAGATTCAGTTGAAGGAAGA**T**GCAGAGAACAATT | SDM (QCL) |
| DES_E203D_rev | AATTGTTCTCTGC**A**TCTTCCTTCAACTGAATCTCCTCCT | SDM (QCL) |
| DES_A204T_for | GGAGATTCAGTTGAAGGAAGAA**ACG**GAGAACAATTTGGCTGCC | SDM (QCL) |
| DES_A204T_rev | GGCAGCCAAATTGTTCTC**CGT**TTCTTCCTTCAACTGAATCTCC | SDM (QCL) |
| DES_A204S_for | GGAGGAGATTCAGTTGAAGGAAGAA**T**CAGAGAACAATTTG | SDM (QCL) |
| DES_A204S_rev | CAAATTGTTCTCTG**A**TTCTTCCTTCAACTGAATCTCCTCC | SDM (QCL) |
| DES_E205D_for | GTTGAAGGAAGAAGCAGA**T**AACAATTTGGCTGCCTTC | SDM (QCL) |
| DES_E205D_rev | GAAGGCAGCCAAATTGTT**A**TCTGCTTCTTCCTTCAAC | SDM (QCL) |
| DES_L208W_for | GAAGGAAGAAGCAGAGAACAATT**G**GGCTGCCTTCCG | SDM (QCL) |
| DES_L208W_rev | CGGAAGGCAGCC**C**AATTGTTCTCTGCTTCTTCCTTC | SDM (QCL) |
| DES_L208S_for | GAAGGAAGAAGCAGAGAACAATT**C**GGCTGCCTTCCG | SDM (QCL) |
| DES_L208S_rev | CGGAAGGCAGCC**G**AATTGTTCTCTGCTTCTTCCTTC | SDM (QCL) |
| DES_A209P_for | AGAAGCAGAGAACAATTTG**C**CTGCCTTCCGAGC | SDM (QCL) |
| DES_A209P_rev | GCTCGGAAGGCAG**G**CAAATTGTTCTCTGCTTCT | SDM (QCL) |
| DES_A210V_for | GAGAACAATTTGGCTG**T**CTTCCGAGCGGACGTG | SDM (QCL) |
| DES_A210V_rev | CACGTCCGCTCGGAAG**A**CAGCCAAATTGTTCTC | SDM (QCL) |
| DES_R212Q_for | TTTGGCTGCCTTCC**A**AGCGGACGTGGATG | SDM (QCL) |
| DES_R212Q_rev | CATCCACGTCCGCT**T**GGAAGGCAGCCAAA | SDM (QCL) |
| DES_A213T_for | AATTTGGCTGCCTTCCGA**A**CGGACGTGGATG | SDM (QCL) |
| DES_A213T_rev | CATCCACGTCCG**T**TCGGAAGGCAGCCAAATT | SDM (QCL) |
| DES_A213V_for | GCTGCCTTCCGAG**T**GGACGTGGATGCA | SDM (QCL) |
| DES_A213V_rev | TGCATCCACGTCC**A**CTCGGAAGGCAGC | SDM (QCL) |
| DES_D214Y_for | GCTGCCTTCCGAGCG**TAT**GTGGATGCAGCTACT | SDM (QCL) |
| DES_D214Y_rev | AGTAGCTGCATCCAC**ATA**CGCTCGGAAGGC AGC | SDM (QCL) |
| DES_V215L_for | CCTTCCGAGCGGAC**T**TGGATGCAGCTACT | SDM (QCL) |
| DES_V215L_rev | AGTAGCTGCATCCA**A**GTCCGCTCGGAAGG | SDM (QCL) |
| DES_V215M_for | CCTTCCGAGCGGAC**A**TGGATGCAGCTACT | SDM (QCL) |
| DES_V215M_rev | AGTAGCTGCATCCA**T**GTCCGCTCGGAAGG | SDM (QCL) |
| DES_T219I_for | CGGACGTGGATGCAGCTA**T**TCTAGCTCGCATTG | SDM (QCL) |
| DES_T219I_rev | CAATGCGAGCTAGA**A**TAGCTGCATCCACGT CCG | SDM (QCL) |
| DES_L220V_for | GTGGATGCAGCTACT**G**TAGCTCGCATTGACC | SDM (QCL) |
| DES_L220V_rev | GGTCAATGCGAGCTA**C**AGTAGCTGCATCCAC | SDM (QCL) |
| DES_A221V_for | GATGCAGCTACTCTAG**T**TCGCATTGACCTGGAG | SDM (QCL) |
| DES_A221V_rev | CTCCAGGTCAATGCGA**A**CTAGAGTAGCTGC ATC | SDM (QCL) |
| DES_R222G_for | GGATGCAGCTACTCTAGCT**G**GCATTGACCTG | SDM (QCL) |
| DES_R222G_rev | CAGGTCAATGC**C**AGCTAGAGTAGCTGCATCC | SDM (QCL) |
| DES_R222C_for | GTGGATGCAGCTACTCTAGCT**T**GCATTGACCTGG | SDM (QCL) |
| DES_R222C_rev | CCAGGTCAATGC**A**AGCTAGAGTAGCTGCATCCAC | SDM (QCL) |
| DES_R222H_for | CAGCTACTCTAGCTC**A**CATTGACCTGGAGCG | SDM (QCL) |
| DES_R222H_rev | CGCTCCAGGTCAATG**T**GAGCTAGAGTAGCTG | SDM (QCL) |
| DES_D224H_for | AGCTACTCTAGCTCGCATT**C**ACCTGGAGCG | SDM (QCL) |
| DES_D224H_rev | CGCTCCAGGT**G**AATGCGAGCTAGAGTAGCT | SDM (QCL) |
| DES_R227C_for | CTCGCATTGACCTGGAG**T**GCAGAATTGAATCTCTC | SDM (QCL) |
| DES_R227C_rev | GAGAGATTCAATTCTGC**A**CTCCAGGTCAATGCGAG | SDM (QCL) |
| DES_R227L_for | CGCATTGACCTGGAGC**T**CAGAATTGAATCTCTCA | SDM (QCL) |
| DES_R227L_rev | TGAGAGATTCAATTCTG**A**GCTCCAGGTCAATGCG | SDM (QCL) |
| DES_R227H_for | CGCATTGACCTGGAGC**A**CAGAATTGAATCTCTCA | SDM (QCL) |
| DES_R227H_rev | TGAGAGATTCAATTCTG**T**GCTCCAGGTCAATGCG | SDM (QCL) |
| DES_L232F_for | ACCTGGAGCGCAGAATTGAATCT**T**TCAACGAGGAGA | SDM (QCL) |
| DES_L232F_rev | TCTCCTCGTTGA**A**AGATTCAATTCTGCGCTCCA GGT | SDM (QCL) |
| DES_E234K_for | CAGAATTGAATCTCTCAAC**A**AGGAGATCGCGTTCCTTAA | SDM (QCL) |
| DES_E234K_rev | TTAAGGAACGCGATCTCCTTGT**T**GAGAGATTCAATTCTG | SDM (QCL) |
| DES_A237S_for | CTCTCAACGAGGAGATC**T**CGTTCCTTAAGAAAGTG | SDM (QCL) |
| DES_A237S_rev | CACTTTCTTAAGGAACG**A**GATCTCCTCGTTGAGAG | SDM (QCL) |
| DES_A237T_for | CTCTCAACGAGGAGATC**A**CGTTCCTTAAGAAAGTG | SDM (QCL) |
| DES_A237T_rev | CACTTTCTTAAGGAACG**T**GATCTCCTCGTTGAGAG | SDM (QCL) |
| DES_A237V_for | TCTCAACGAGGAGATCG**T**GTTCCTTAAGAAAGTGC | SDM (QCL) |
| DES_A237V_rev | GCACTTTCTTAAGGAAC**A**CGATCTCCTCGTTGAGA | SDM (QCL) |
| DES_L239F_for | AACGAGGAGATCGCGTTC**T**TTAAGAAAGTGCATGAAG | SDM (QCL) |
| DES_L239F_rev | CTTCATGCACTTTCTTAA**A**GAACGCGATCTCCTCGTT | SDM (QCL) |
| DES_K240E_for | CGAGGAGATCGCGTTCCTT**G**AGAAAGTGCATGAAG | SDM (QCL) |
| DES_K240E_rev | CTTCATGCACTTTCT**C**AAGGAACGCGATCTCCTCG | SDM (QCL) |
| DES_V242E_for | CGCGTTCCTTAAGAAAG**A**GCATGAAGAGGAGATCC | SDM (QCL) |
| DES_V242E_rev | GGATCTCCTCTTCATGC**T**CTTTCTTAAGGAACGCG | SDM (QCL) |
| DES_H243Y_for | GCGTTCCTTAAGAAAGTG**T**ATGAAGAGGAGATCCGTG | SDM (QCL) |
| DES_H243Y_rev | CACGGATCTCCTCTTCAT**A**CACTTTCTTAAGGAACGC | SDM (QCL) |
| DES_H243R_for | GTTCCTTAAGAAAGTGC**G**TGAAGAGGAGATCCGTG | SDM (QCL) |
| DES_H243R_rev | CACGGATCTCCTCTTCA**C**GCACTTTCTTAAGGAAC | SDM (QCL) |
| DES_E246D_for | AAAGTGCATGAAGAGGA**T**ATCCGTGAGTTGCAGGC | SDM (QCL) |
| DES_E246D_rev | GCCTGCAACTCACGGAT**A**TCCTCTTCATGCACTTT | SDM (QCL) |
| DES_E246K_for | AGAAAGTGCATGAAGAG**A**AGATCCGTGAGTTGCAG | SDM (QCL) |
| DES_E246K_rev | CTGCAACTCACGGATCT**T**CTCTTCATGCACTTTCT | SDM (QCL) |
| DES_R248H_for | CATGAAGAGGAGATCC**A**TGAGTTGCAGGCTCAG | SDM (QCL) |
| DES_R248H_rev | CTGAGCCTGCAACTCA**T**GGATCTCCTCTTCATG | SDM (QCL) |
| DES_R248C_for | GCATGAAGAGGAGATC**T**GTGAGTTGCAGGCTCA | SDM (QCL) |
| DES_R248C_rev | TGAGCCTGCAACTCAC**A**GATCTCCTCTTCATGC | SDM (QCL) |
| DES_R248L_for | CATGAAGAGGAGATCC**T**TGAGTTGCAGGCTCAG | SDM (QCL) |
| DES_R248L_rev | CTGAGCCTGCAACTCA**A**GGATCTCCTCTTCATG | SDM (QCL) |
| DES_R248A_for | GCATGAAGAGGAGATC**GC**TGAGTTGCAGGCTCAG | SDM (QCL) |
| DES_R248A_rev | CTGAGCCTGCAACTCA**GC**GATCTCCTCTTCATGC | SDM (QCL) |
| DES_R248S_for | AGAGGAGATC**AGC**GAGTTGCAGGC | SDM (Q5) |
| DES_R248S_rev | TCATGCACTTTCTTAAGGAAC | SDM (Q5) |
| DES_E249A_for | AGAGGAGATCCGTG**C**GTTGCAGGCTCAGC | SDM (QCL) |
| DES_E249A_rev | GCTGAGCCTGCAAC**G**CACGGATCTCCTCT | SDM (QCL) |
| DES_A152P_for | GACGCGAGTG**CCG**GAGCTCTACGAGGAGG | SDM (Q5) |
| DES_A152P_rev | GGCTCGCGGCCCTTGAGC | SDM (Q5) |
| DES_E153P_for | GACGCGAGTGGCC**CC**GCTCTACGAGGAG | SDM (QCL) |
| DES_E153P_rev | CTCCTCGTAGAGC**GG**GGCCACTCGCGTC | SDM (QCL) |
| DES_Y155P_for | GGCCGAGCTC**CCG**GAGGAGGAGCTGCGG | SDM (Q5) |
| DES_Y155P_rev | ACTCGCGTCGGCTCGCGG | SDM (Q5) |
| DES_E156P_for | GGCCGAGCTCTAC**CC**GGAGGAGCTGCGG | SDM (QCL) |
| DES_E156P_rev | CCGCAGCTCCTCC**GG**GTAGAGCTCGGCC | SDM (QCL) |
| DES_E157P_for | CGAGCTCTACGAG**CC**GGAGCTGCGGGAG | SDM (QCL) |
| DES_E157P_rev | CTCCCGCAGCTCC**GG**CTCGTAGAGCTCG | SDM (QCL) |
| DES_E158P_for | CTACGAGGAG**CC**GCTGCGGGAGCTGC | SDM (Q5) |
| DES_E158P_rev | AGCTCGGCCACTCGCGTC | SDM (Q5) |
| DES_R160P_for | GGAGGAGCTGC**C**GGAGCTGCGGC | SDM (Q5) |
| DES_R160P_rev | TCGTAGAGCTCGGCCACTCG | SDM (Q5) |
| DES_E161P_for | GGAGCTGCGG**CC**GCTGCGGCGCC | SDM (Q5) |
| DES_E161P_rev | TCCTCGTAGAGCTCGGCC | SDM (Q5) |
| DES_L162P_for | GCTGCGGGAGC**C**GCGGCGCCAGG | SDM (Q5) |
| DES_L162P_rev | TCCTCCTCGTAGAGCTCGGCCACTCG | SDM (Q5) |
| DES_R164P_for | GGAGCTGCGGC**C**CCAGGTGGAGG | SDM (QCL) |
| DES_R164P_rev | CCTCCACCTGG**G**GCCGCAGCTCC | SDM (QCL) |
| DES_Q165P_for | GCTGCGGCGCC**C**GGTGGAGGTGC | SDM (QCL) |
| DES_Q165P_rev | GCACCTCCACC**G**GGCGCCGCAGC | SDM (QCL) |
| DES_V166P_for | GCTGCGGCGCCAG**CC**GGAGGTGCTCACT | SDM (QCL) |
| DES_V166P_rev | AGTGAGCACCTCC**GG**CTGGCGCCGCAGC | SDM (QCL) |
| DES_E167P_for | GCGGCGCCAGGTG**CC**GGTGCTCACTAAC | SDM (QCL) |
| DES_E167P_rev | GTTAGTGAGCACC**GG**CACCTGGCGCCGC | SDM (QCL) |
| DES_V168P_for | GGCGCCAGGTGGAG**CC**GCTCACTAACCAGC | SDM (QCL) |
| DES_V168P_rev | GCTGGTTAGTGAGC**GG**CTCCACCTGGCGCC | SDM (QCL) |
| DES_L169P_for | CAGGTGGAGGTGC**C**CACTAACCAGCGC | SDM (QCL) |
| DES_L169P_rev | GCGCTGGTTAGTG**G**GCACCTCCACCTG | SDM (QCL) |
| DES_T170P_for | GGTGGAGGTGCTC**C**CTAACCAGCGCGC | SDM (QCL) |
| DES_T170P_rev | GCGCGCTGGTTAG**G**GAGCACCTCCACC | SDM (QCL) |
| DES_N171P_for | CCAGGTGGAGGTGCTCACT**CC**CCAGCGCGCG | SDM (QCL) |
| DES_N171P_rev | CGCGCGCTGG**GG**AGTGAGCACCTCCACCTGG | SDM (QCL) |
| DES_Q172P_for | GCTCACTAACC**C**GCGCGCGCGCG | SDM (QCL) |
| DES_Q172P_rev | CGCGCGCGCGC**G**GGTTAGTGAGC | SDM (QCL) |
| DES_R173P_for | CACTAACCAGC**C**CGCGCGCGTCG | SDM (QCL) |
| DES_R173P_rev | CGACGCGCGCG**G**GCTGGTTAGTG | SDM (QCL) |
| DES_A174P_for | CTAACCAGCGC**C**CGCGCGTCGAC | SDM (QCL) |
| DES_A174P_rev | GTCGACGCGCG**G**GCGCTGGTTAG | SDM (QCL) |
| DES_R175P_for | CCAGCGCGCGC**CG**GTCGACGTCG | SDM (Q5) |
| DES_R175P_rev | TTAGTGAGCACCTCCACCTG | SDM (Q5) |
| DES_V176P_for | CAGCGCGCGCGC**CC**CGACGTCGAGCG | SDM (QCL) |
| DES_V176P_rev | CGCTCGACGTCG**GG**GCGCGCGCGCTG | SDM (QCL) |
| DES_D177P_for | GCGCGCGCGTC**CC**CGTCGAGCGCG | SDM (QCL) |
| DES_D177P_rev | CGCGCTCGACG**GG**GACGCGCGCGC | SDM (QCL) |
| DES_V178P_for | CGCGCGCGTCGAC**CC**CGAGCGCGACAAC | SDM (QCL) |
| DES_V178P_rev | GTTGTCGCGCTCG**GG**GTCGACGCGCGCG | SDM (QCL) |
| DES_E179P_for | GCGCGTCGACGTC**CC**GCGCGACAACCTG | SDM (QCL) |
| DES_E179P_rev | CAGGTTGTCGCGCG**GG**ACGTCGACGCGC | SDM (QCL) |
| DES_R180P_for | GTCGACGTCGAGC**C**CGACAACCTGCTC | SDM (QCL) |
| DES_R180P_rev | GAGCAGGTTGTCG**G**GCTCGACGTCGAC | SDM (QCL) |
| DES_D181P_for | CGACGTCGAGCGC**CC**CAACCTGCTCGAC | SDM (QCL) |
| DES_D181P_rev | GTCGAGCAGGTTG**GG**GCGCTCGACGTCG | SDM (QCL) |
| DES_N182P_for | CGTCGAGCGCGAC**CC**CCTGCTCGACGAC | SDM (QCL) |
| DES_N182P_rev | GTCGTCGAGCAG**GG**GGTCGCGCTCGACG | SDM (QCL) |
| DES_L183P_for | GAGCGCGACAACC**C**GCTCGACGACCTG | SDM (QCL) |
| DES_L183P_rev | CAGGTCGTCGAGCGG**G**TTGTCGCGCTC | SDM (QCL) |
| DES_L184P_for | CGCGACAACCTGC**C**CGACGACCTGCAG | SDM (QCL) |
| DES_L184P_rev | CTGCAGGTCGTCG**G**GCAGGTTGTCGCG | SDM (QCL) |
| DES_D185P_for | CGACAACCTGCTC**CC**CGACCTGCAGCGG | SDM (QCL) |
| DES_D185P_rev | CCGCTGCAGGTCG**GG**GAGCAGGTTGTCG | SDM (QCL) |
| DES_D186P_for | CAACCTGCTCGAC**CC**CCTGCAGCGGCTC | SDM (QCL) |
| DES_D186P_rev | GAGCCGCTGCAG**GG**GGTCGAGCAGGTTG | SDM (QCL) |
| DES_Q188P_for | TCGACGACCTGC**C**GCGGCTCAAGGC | SDM (QCL) |
| DES_Q188P_rev | GCCTTGAGCCGCG**G**CAGGTCGTCGA | SDM (QCL) |
| DES_R189P_for | GACGACCTGCAGC**C**GCTCAAGGC | Overlap-PCR |
| DES_R189P_rev | GCCTTGAGC**G**GCTGCAGGTCGTC | Overlap-PCR |
| DES_L190P_for | GACCTGCAGCGGC**C**CAAGGCCAAGCTG | SDM (QCL) |
| DES_L190P_rev | CAGCTTGGCCTTG**G**GCCGCTGCAGGTC | SDM (QCL) |
| DES_K191P_for | GCAGCGGCTC**CC**GGCCAAGCTGC | SDM (Q5) |
| DES_K191P_rev | AGGTCGTCGAGCAGGTTG | SDM (Q5) |
| DES_A192P_for | CAGCGGCTCAAG**C**CCAAGCTGCAGG | SDM (QCL) |
| DES_A192P_rev | CCTGCAGCTTGG**G**CTTGAGCCGCTG | SDM (QCL) |
| DES_K193P_for | GCTCAAGGCC**CC**GCTGCAGGAGGAG | SDM (Q5) |
| DES_K193P_rev | CGCTGCAGGTCGTCGAGC | SDM (Q5) |
| DES_L194P_for | GCTCAAGGCCAAGC**C**GCAGGAGGAGATTC | SDM (QCL) |
| DES_L194P_rev | GAATCTCCTCCTGC**G**GCTTGGCCTTGAGC | SDM (QCL) |
| DES_Q195P_for | CTCAAGGCCAAGCTGC**C**GGAGGAGATTCAGTTG | SDM (QCL) |
| DES_Q195P_rev | CAACTGAATCTCCTCC**G**GCAGCTTGGCCTTGAG | SDM (QCL) |
| DES_E196P_for | TCAAGGCCAAGCTGCAG**CC**GGAGATTCAGTTGAAGG | SDM (QCL) |
| DES_E196P_rev | CCTTCAACTGAATCTCC**GG**CTGCAGCTTGGCCTTGA | SDM (QCL) |
| DES_E197P_for | AGGCCAAGCTGCAGGAG**CC**GATTCAGTTGAAGGAAG | SDM (QCL) |
| DES_E197P_rev | CTTCCTTCAACTGAATC**GG**CTCCTGCAGCTTGGCCT | SDM (QCL) |
| DES_I198P_for | GCCAAGCTGCAGGAGGAG**CC**TCAGTTGAAGGAAGAAGC | SDM (QCL) |
| DES_I198P_rev | GCTTCTTCCTTCAACTGA**GG**CTCCTCCTGCAGCTTGGC | SDM (QCL) |
| DES_Q199P_for | GGAGGAGATTC**C**GTTGAAGGAAGAAGC | SDM (Q5) |
| DES_Q199P_rev | TGCAGCTTGGCCTTGAGC | SDM (Q5) |
| DES_L200P_for | GGAGATTCAG**CC**GAAGGAAGAAGCAGAGAAC | SDM (Q5) |
| DES_L200P_rev | TCCTGCAGCTTGGCCTTG | SDM (Q5) |
| DES_K201P_for | GCAGGAGGAGATTCAGTTG**CC**GGAAGAAGCAGAGAACAAT | SDM (QCL) |
| DES_K201P_rev | ATTGTTCTCTGCTTCTTCC**GG**CAACTGAATCTCCTCCTGC | SDM (QCL) |
| DES_E202P_for | TCAGTTGAAG**CCG**GAAGCAGAGAACAATTTGG | SDM (Q5) |
| DES_E202P_rev | ATCTCCTCCTGCAGCTTG | SDM (Q5) |
| DES_E203P_for | GTTGAAGGAA**CCG**GCAGAGAACAATTTGGCTGC | SDM (Q5) |
| DES_E203P_rev | TGAATCTCCTCCTGCAGC | SDM (Q5) |
| DES_A204P_for | GGAGGAGATTCAGTTGAAGGAAGAA**C**CAGAGAACAATTTG | SDM (QCL) |
| DES_A204P_rev | CAAATTGTTCTCTG**G**TTCTTCCTTCAACTGAATCTCCTCC | SDM (QCL) |
| DES_E205P_for | TCAGTTGAAGGAAGAAGCA**CC**GAACAATTTGGCTGCCTTC | SDM (QCL) |
| DES_E205P_rev | GAAGGCAGCCAAATTGTTC**GG**TGCTTCTTCCTTCAACTGA | SDM (QCL) |
| DES_N206P_for | GTTGAAGGAAGAAGCAGAG**CC**CAATTTGGCTGCCTTCCGA | SDM (QCL) |
| DES_N206P_rev | TCGGAAGGCAGCCAAATTG**GG**CTCTGCTTCTTCCTTCAAC | SDM (QCL) |
| DES_N207P_for | AGCAGAGAAC**CCG**TTGGCTGCCTTCC | SDM (Q5) |
| DES_N207P_rev | TCTTCCTTCAACTGAATCTCC | SDM (Q5) |
| DES_L208P_for | GTTGAAGGAAGAAGCAGAGAACAAT**CC**GGCTGCCTTCCGA | SDM (QCL) |
| DES_L208P_rev | TCGGAAGGCAGCC**GG**ATTGTTCTCTGCTTCTTCCTTCAAC | SDM (QCL) |
| DES_A209P_for | GAACAATTTG**CCG**GCCTTCCGAGC | SDM (QCL) |
| DES_A209P_rev | TCTGCTTCTTCCTTCAACTG | SDM (QCL) |
| DES_A210P_for | CAATTTGGCT**CCG**TTCCGAGCGG | SDM (Q5) |
| DES_A210P_rev | TTCTCTGCTTCTTCCTTCAAC | SDM (Q5) |
| DES_F211P_for | GAACAATTTGGCTGCC**CCG**CGAGCGGACGTG | Overlap-PCR |
| DES_F211P_rev | CACGTCCGCTCG**CGG**GGCAGCCAAATTGTT | Overlap-PCR |
| DES_R212P_for | GGCTGCCTTCC**C**AGCGGACGTGG | SDM (QCL) |
| DES_R212P_rev | CCACGTCCGCT**G**GGAAGGCAGC | SDM (QCL) |
| DES_A213P_for | TGCCTTCCGA**C**CGGACGTGGATG | SDM (Q5) |
| DES_A213P_rev | GCCAAATTGTTCTCTGCTTCTTCC | SDM (Q5) |
| DES_D214P_for | CTTCCGAGCG**CCG**GTGGATGCAGC | SDM (Q5) |
| DES_D214P_rev | GCAGCCAAATTGTTCTCTG | SDM (Q5) |
| DES_V215P_for | CCTTCCGAGCGGAC**CC**GGATGCAGCTACTC | SDM (QCL) |
| DES_V215P_rev | GAGTAGCTGCATCC**GG**GTCCGCTCGGAAGG | SDM (QCL) |
| DES_D216P_for | AGCGGACGTG**CCG**GCAGCTACTC | SDM (Q5) |
| DES_D216P_rev | CGGAAGGCAGCCAAATTG | SDM (Q5) |
| DES_A217P_for | GGACGTGGAT**CCG**GCTACTCTAGC | SDM (Q5) |
| DES_A217P_rev | GCTCGGAAGGCAGCCAAA | SDM (Q5) |
| DES_A218P_for | CGTGGATGCA**CCG**ACTCTAGCTCGCATTGAC | SDM (Q5) |
| DES_A218P_rev | TCCGCTCGGAAGGCAGCC | SDM (Q5) |
| DES_T219P_for | GACGTGGATGCAGCT**C**CTCTAGCTCGCATTG | SDM (QCL) |
| DES_T219P_rev | CAATGCGAGCTAGAG**G**AGCTGCATCCACGTC | SDM (QCL) |
| DES_L220P_for | TGCAGCTACTC**CG**GCTCGCATTGACC | SDM (Q5) |
| DES_L220P_rev | TCCACGTCCGCTCGGAAG | SDM (Q5) |
| DES_A221P_for | AGCTACTCTA**CCG**CGCATTGACCTGGAGC | SDM (Q5) |
| DES_A221P_rev | GCATCCACGTCCGCTCGG | SDM (Q5) |
| DES_R222P_for | CAGCTACTCTAGCTC**C**CATTGACCTGGAGC | SDM (QCL) |
| DES_R222P_rev | CGCTCCAGGTCAATG**G**GAGCTAGAGTAGCTG | SDM (QCL) |
| DES_I223P_for | TCTAGCTCGC**CCG**GACCTGGAGCG | SDM (Q5) |
| DES_I223P_rev | GTAGCTGCATCCACGTCC | SDM (Q5) |
| DES_D224P_for | AGCTCGCATT**CCG**CTGGAGCGCA | SDM (Q5) |
| DES_D224P_rev | AGAGTAGCTGCATCCACG | SDM (Q5) |
| DES_L225P_for | TCGCATTGACC**C**GGAGCGCAGAA | SDM (Q5) |
| DES_L225P_rev | GCTAGAGTAGCTGCATCCAC | SDM (Q5) |
| DES_E226P_for | CTAGCTCGCATTGACCTG**CC**GCGCAGAATTGAATCTCT | SDM (QCL) |
| DES_E226P_rev | AGAGATTCAATTCTGCGC**GG**CAGGTCAATGCGAGCTAG | SDM (QCL) |
| DES_R227P_for | TGACCTGGAGC**CG**AGAATTGAATCTC | SDM (Q5) |
| DES_R227P_rev | ATGCGAGCTAGAGTAGCT | SDM (Q5) |
| DES_R228P_for | CCTGGAGCGC**CCG**ATTGAATCTCTC | SDM (Q5) |
| DES_R228P_rev | TCAATGCGAGCTAGAGTAG | SDM (Q5) |
| DES_I229P_for | GCATTGACCTGGAGCGCAGA**CC**TGAATCTCTCAACGAG | SDM (QCL) |
| DES_I229P_rev | CTCGTTGAGAGATTCA**GG**TCTGCGCTCCAGGTCAATGC | SDM (QCL) |
| DES_E230P_for | GCGCAGAATT**CCG**TCTCTCAACGAGGAGATC | SDM (Q5) |
| DES_E230P_rev | TCCAGGTCAATGCGAGCT | SDM (Q5) |
| DES_S231P_for | GCGCAGAATTGAA**C**CTCTCAACGAGGAGATC | Overlap-PCR |
| DES_S231P_rev | GATCTCCTCGTTGAGAG**G**TTCAATTCTGCGC | Overlap-PCR |
| DES_L232P_for | AATTGAATCTC**CG**AACGAGGAGATCGCG | SDM (Q5) |
| DES_L232P_rev | CTGCGCTCCAGGTCAATG | SDM (Q5) |
| DES_N233P_for | CGCAGAATTGAATCTCTC**CC**CGAGGAGATCGCGTTCCT | SDM (QCL) |
| DES_N233P_rev | AGGAACGCGATCTCCTCG**GG**GAGAGATTCAATTCTGCG | SDM (QCL) |
| DES_E234P_for | ATCTCTCAAC**CC**GGAGATCGCGTTCC | SDM (Q5) |
| DES_E234P_rev | TCAATTCTGCGCTCCAGG | SDM (Q5) |
| DES_E235P_for | TCTCAACGAG**CC**GATCGCGTTCC | SDM (Q5) |
| DES_E235P_rev | GATTCAATTCTGCGCTCC | SDM (Q5) |
| DES_I236P_for | TGAATCTCTCAACGAGGAG**CC**CGCGTTCCTTAAGAAAGTG | SDM (QCL) |
| DES_I236P_rev | CACTTTCTTAAGGAACGCG**GG**CTCCTCGTTGAGAGATTCA | SDM (QCL) |
| DES_A237P_for | CGAGGAGATC**C**CGTTCCTTAAGAAAG | SDM (Q5) |
| DES_A237P_rev | TTGAGAGATTCAATTCTGCGC | SDM (Q5) |
| DES_F238P_for | GGAGATCGCG**CCG**CTTAAGAAAGTGCATG | SDM (Q5) |
| DES_F238P_rev | TCGTTGAGAGATTCAATTCTGC | SDM (Q5) |
| DES_L239P_for | GATCGCGTTCC**CG**AAGAAAGTGC | SDM (Q5) |
| DES_L239P_rev | TCCTCGTTGAGAGATTCAATTC | SDM (Q5) |
| DES_K240P_for | CGAGGAGATCGCGTTCCTT**CC**GAAAGTGCATGAAGAGGAG | SDM (QCL) |
| DES_K240P_rev | CTCCTCTTCATGCACTTTC**GG**AAGGAACGCGATCTCCTCG | SDM (QCL) |
| DES_K241P_for | GTTCCTTAAG**CCG**GTGCATGAAGAGGAGATC | SDM (Q5) |
| DES_K241P_rev | GCGATCTCCTCGTTGAGA | SDM (Q5) |
| DES_V242P_for | CCTTAAGAAA**CC**GCATGAAGAGGAGATCCGTG | SDM (Q5) |
| DES_V242P_rev | AACGCGATCTCCTCGTTG | SDM (Q5) |
| DES_H243P_for | GTTCCTTAAGAAAGTGC**C**TGAAGAGGAGATCCGTG | SDM (QCL) |
| DES_H243P_rev | CACGGATCTCCTCTTCA**G**GCACTTTCTTAAGGAAC | SDM (QCL) |
| DES_E244P_for | GAAAGTGCAT**CCG**GAGGAGATCCGTG | SDM (Q5) |
| DES_E244P_rev | TTAAGGAACGCGATCTCC | SDM (Q5) |
| DES_E245P_for | AGTGCATGAA**CC**GGAGATCCGTGAGTTGC | SDM (Q5) |
| DES_E245P_rev | TTCTTAAGGAACGCGATCTC | SDM (Q5) |
| DES_E246P_for | GCATGAAGAG**CC**GATCCGTGAGTTGC | SDM (Q5) |
| DES_E246P_rev | ACTTTCTTAAGGAACGCGATC | SDM (Q5) |
| DES_I247P_for | AAGTGCATGAAGAGGAG**CC**CCGTGAGTTGCAGGCTC | SDM (QCL) |
| DES_I247P_rev | GAGCCTGCAACTCACGG**GG**CTCCTCTTCATGCACTT | SDM (QCL) |
| DES_R248P_for | AGAGGAGATCC**CG**GAGTTGCAGGC | SDM (Q5) |
| DES_R248P_rev | TCATGCACTTTCTTAAGGAAC | SDM (Q5) |
| DES_E249P_for | GGAGATCCGT**CC**GTTGCAGGCTC | SDM (Q5) |
| DES_E249P_rev | TCTTCATGCACTTTCTTAAGG | SDM (Q5) |
| DES_L250P_for | GATCCGTGAG**CC**GCAGGCTCAGC | SDM (Q5) |
| DES_L250P_rev | TCCTCTTCATGCACTTTCTTAAG | SDM (Q5) |
| DES_Q251P_for | GATCCGTGAGTTGC**C**GGCTCAGCTTCAGG | SDM (QCL) |
| DES_Q251P_rev | CCTGAAGCTGAGCC**G**GCAACTCACGGATC | SDM (QCL) |
| DES_A252P_for | GTGAGTTGCAG**C**CTCAGCTTCAGGAAC | Overlap-PCR |
| DES_A252P_rev | GTTCCTGAAGCTGAG**G**CTGCAACTCAC | Overlap-PCR |
| DES_Q253P_for | GTTGCAGGCTC**C**GCTTCAGGAAC | SDM (Q5) |
| DES_Q253P_rev | TCACGGATCTCCTCTTCATG | SDM (Q5) |
| DES_L254P_for | GTTGCAGGCTCAGC**C**TCAGGAACAGCAGG | SDM (QCL) |
| DES_L254P_rev | CCTGCTGTTCCTGAGGCTGAGCCTGCAAC | SDM (QCL) |
| DES_Q255P_for | GGCTCAGCTTC**C**GGAACAGCAGG | SDM (Q5) |
| DES_Q255P_rev | TGCAACTCACGGATCTCC | SDM (Q5) |
| DES_E256P_for | TCAGCTTCAG**CCG**CAGCAGGTCCAGG | SDM (Q5) |
| DES_E256P_rev | GCCTGCAACTCACGGATC | SDM (Q5) |

SDM=Site Directed Mutagenesis; PCR=Polymerase Chain Reaction. Q5=Q5 Site Directed Mutagenesis Kit (NEB). QCL=Quik Change Lightning Site Directed Mutagenesis Kit (Agilent). Mutated Nucleotides are highlighted in red and bold.

**Table S2.** List of the analyzed cardiomyopathy related genes.

*ABCA1, ABCC8, ABCC9, ABCG5, ABCG8, ACTA1, ACTA2, ACTC1, ACTN2, ACVRL1, AKAP9, ALMS1, ALPK3, ANK2, ANKRD1, APOA1, APOA2, APOA5, APOB, APOC2, APOC3, APOE, AQP1, BAG3, BGN, BMPR2, BRAF, CACNA1C, CACNA2D1, CACNB2, CALM1, CALM2, CALM3, CALR3, CASQ2, CAV3, CDH2, CETP, COL1A1, COL1A2, COL3A1, COL4A1, COL4A5, COL5A1, COL5A2, COX15, CRYAB, CSRP3, CTF1, CTNNA3, CYP27A1, DES, DMD, DOLK, DSC2, DSG2, DSP, DTNA, EFEMP2, EIF2AK4, ELN, EMD, EMILIN1, ENG, EPHX2, EYA4, FBN1, FBN2, FHL1, FHL2, FKRP, FKTN, FLNA, FLNC, FOXE3, FXN, GAA, GATAD1,, GCK, GCKR, GDF2, GHR, GLA, GPD1, GPD1L, GPIHBP1, GSBS/PPP1R17, HCN4, HFE, HNF1A, HNF1B, HNF4A, ILK, INS, JPH2, JUP, KCND3, KCNE1, KCNE2, KCNE3, KCNE5, KCNH2, KCNJ11, KCNJ2, KCNJ5, KCNJ8, KCNK3, KCNQ1, KDR, KLF10, KRAS, LAMA4, LAMP2, LCAT, LDB3, LDLR, LDLRAP1, LIPA, LIPC, LMF1, LMNA, LOX, LPL, LZTR1, MAP2K1, MAP2K2, MAT2A, MEFV, MFAP5, MIB1, MRAS, MTTP, MYBPC3, MYH11, MYH6, MYH7, MYL2, MYL3, MYLK, MYLK2, MYO6, MYOM1, MYOZ2, MYPN, NEBL, NEXN, NKX2-5, NOTCH1, NRAS, OBSCN, PCSK9, PDLIM3, PKP2, PLN, PPCS, PPP1CB, PRDM16, PRKAG2, PRKG1, PSEN1, PSEN2, PTPN11, RAF1, RANGRF, RASA2, RBM20, RIT1, ROBO4, RRAS, RRAS2, RYR2, SAR1B, SCARB1, SCN10A, SCN1B, SCN2B, SCN3B, SCN4B, SCN5A, SDHA, SGCD, SHOC2, SKI, SLC25A4, SLC2A10, SLC4A3, SLMAP, SMAD2, SMAD3, SMAD4, SMAD9, SNTA1, SOS1, SOS2, SOX17, STAP1, TAZ, TBX20, TBX4, TCAP, TECRL, TGFB2, TGFB3, TGFBR1, TGFBR2, THSD4, TJP1, TMEM43, TMPO, TNNC1, TNNI3, TNNI3K, TNNT2, TNXB, TPM1, TRDN, TRIM63, TRPM4, TTN, TTR, TXNRD2, VCL, ZBTB17.*

**Table S3.** Overview about the identified coding, non-synonymous genetic variants with a minor allele frequency (MAF) <0.0001 identified by a NGS approach.

| **Gene** | **Variant** | **Kind of Mutation** | **Genotype** | **MAF^1^** | **ACMG Criteria** | **ACMG Classification** |
| --- | --- | --- | --- | --- | --- | --- |
| *DES* | 2-219419022-T-C | Missense variant | heterozygous | 0.0000006479 | PS3, PM2, PP3, PP2 | Likely Pathogenic |
| *TTN* | 2-178537766-G-T | Nonsense variant | heterozygous | 0 | PVS1, PM2 | Likely Pathogenic |
| *LDB3* | 10-86679445-G-A | Missense variant | heterozygous | 0.000003717 | PM2 | VUS |
| *ALMS1* | 2-73451980-G-T | Missense variant | heterozygous | 0.00007505 | PM2 | VUS |
| *SOS1* | 2-39013462-C-T | Missense variant and splice region variant | heterozygous | 0.00008736 | PM2, BS2, PB4, BP6 | Likely Benign |

^1^ According to the Genome Aggregation Database, 26^th^ May 2025.

**Table S4.** Overview about the mutation *DES*-p.L187P.

| Name of the variant | | *DES*-p.L187P (NP_001918.3) | |
| --- | --- | --- | --- |
|  |  | *DES*-c.560T>C (NM_001927.4) | |
| Chromosomal position | | 2-219419022-T-C (GRCh38.p14 chr 2) | |
| rs-number | | [rs1248833348](https://varsome.com/variant/hg38/rs1248833348?&annotation-mode=germline) | <https://www.ncbi.nlm.nih.gov/snp/?term=rs1248833348> |
| Affected protein domain | | 1B | |
| ACMG criteria | | PS3 (moderate), PP3 (moderate),PP2 (supporting) and PM2 (supporting) | |
| Database Information | ClinGen (Gene Disease Validity), 26^th^ May 2025 | Dilated Cardiomyopathy – Definitive  Arrhythmogenic Right Ventricular Cardiomyopathy - Moderate | <https://search.clinicalgenome.org/kb/genes/HGNC:2770> |
|  | gnomAD v4.1.0 (Minor Allele Frequency), 26^th^ May 2025 | 6.479e^-7^ | <https://gnomad.broadinstitute.org/variant/2-219419022-T-C?dataset=gnomad_r4> |
|  | RGC Million Exome Variant Brower (Alternate Allele Frequency), 26^th^ May 2025 | Not listed | <https://rgc-research.regeneron.com/me/gene/DES> |
|  | ClinVar, 26^th^ May 2025 | Uncertain Significane | <https://www.ncbi.nlm.nih.gov/clinvar/variation/1678019/> |
| PhyloP100way (Conservation Score) | | 7.794 | |
| In-Silico Predictors  (Meta Scores) | BayesDel addAF | Pathogenic Strong (0.4556) | |
|  | MetaRNN | Pathogenic Strong (0.9429) | |
|  | REVEL | Pathogenic Moderate (0.922) | |
|  | BayesDel noAF | Pathogenic Moderate (0.4951) | |
|  | MetaLR | Pathogenic Supporting (0.8442) | |
|  | MetaSVM | Pathogenic Moderate (0.9389) | |
| In Silico Predictors (Individual Predictions) | AlphaMissense | Pathogenic Strong (0.9963) | |
|  | EIGEN | Pathogenic Strong (0.8805) | |
|  | DEOGEN2 | Pathogenic Moderate (0.9176) | |
|  | EIGEN PC | Pathogenic Supporting (0.7712) | |
|  | EVE | Pathogenic Moderate (0.798) | |
|  | FATHMM-XF | Pathogenic Moderate (0.9937) | |
|  | M-CAP | Uncertain (0.2571) | |
|  | Mutation assessor | Pathogenic Moderate (3.71) | |
|  | MutPred | Pathogenic Moderate (0.747) | |
|  | MVP | Pathogenic Moderate (0.9723) | |
|  | FATHMM-MKL | Pathogenic Supporting (0.9792) | |
|  | LIST-S2 | Uncertain (0.9039) | |
|  | PrimateAI | Pathogenic Supporting (0.8133) | |
|  | PROVEAN | Pathogenic Supporting (-6.08) | |
|  | SIFT | Pathogenic Supporting (0) | |
|  | SIFT4G | Pathogenic Supporting (0.002) | |

**Figure S1.** Plasmid Map of pEYFP-N1-DES.

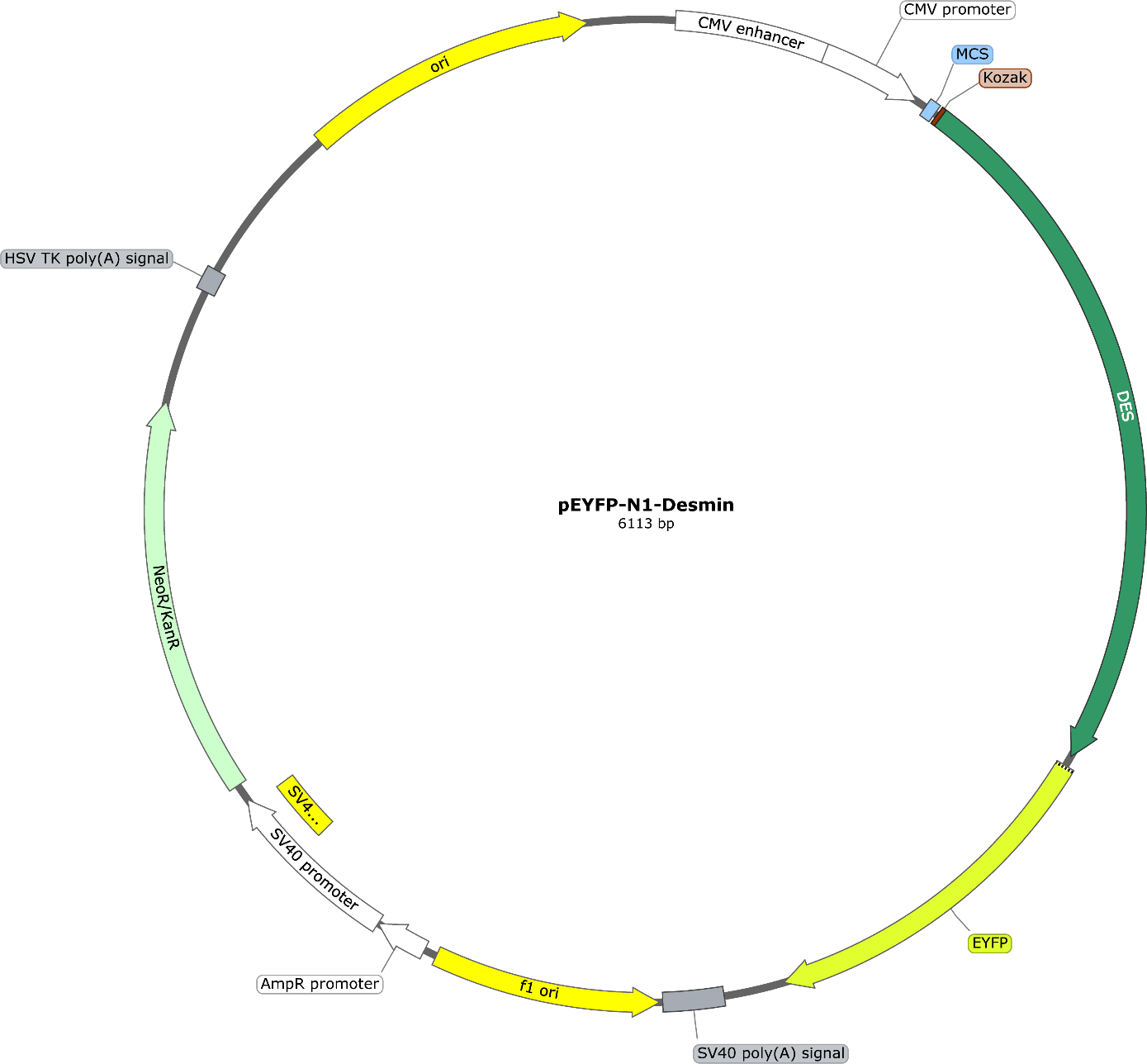

**Figure S2.** Plasmid Map of pmRuby-N1-DES.

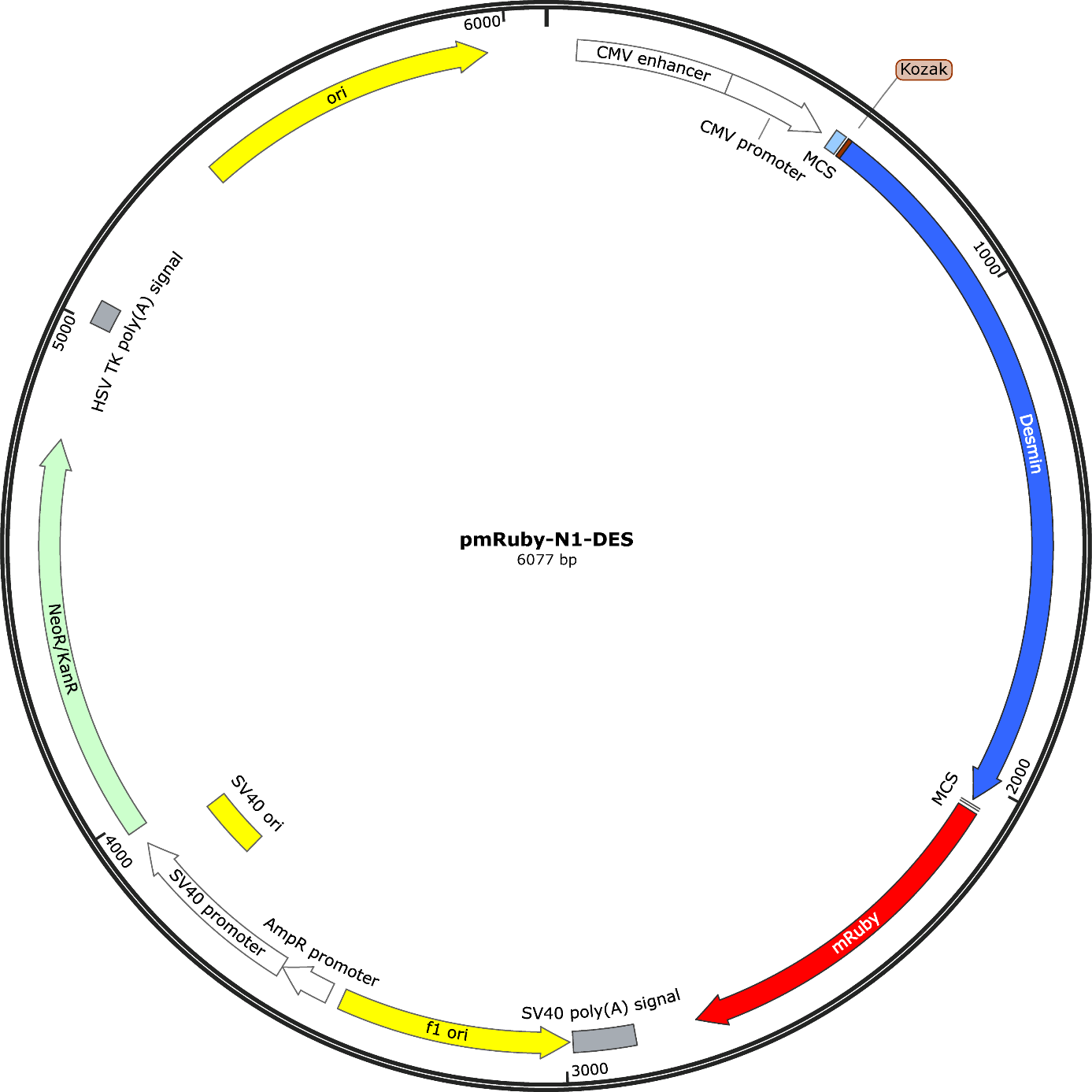

**Figure S3.** Plasmid Map of pET100D-TOPO-DES.

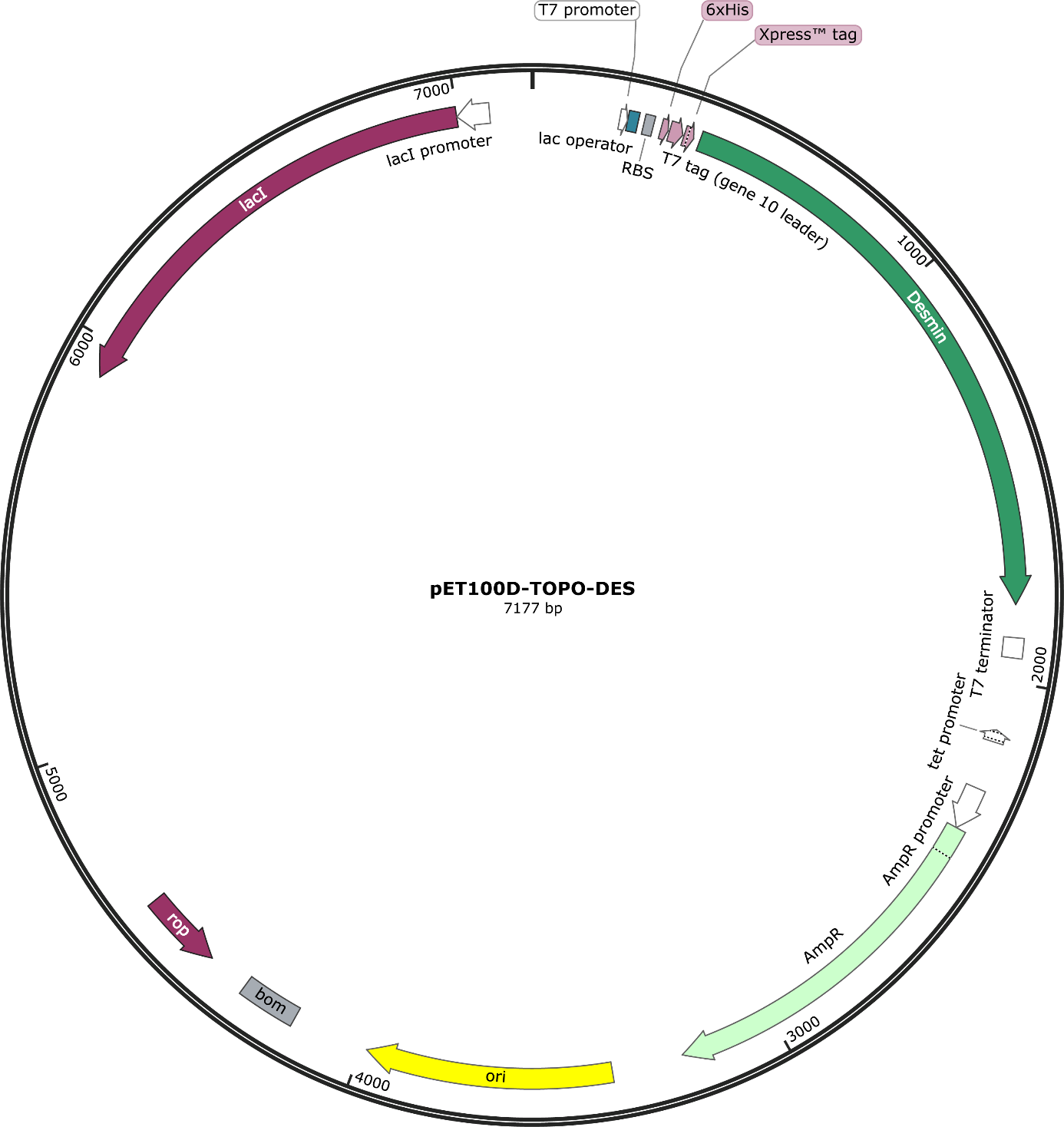

**Figure S4.** Partial electropherograms of the desmin expression constructs (pEYFP-N1-DES) carrying the variants of unknown significance within the 1B domain.

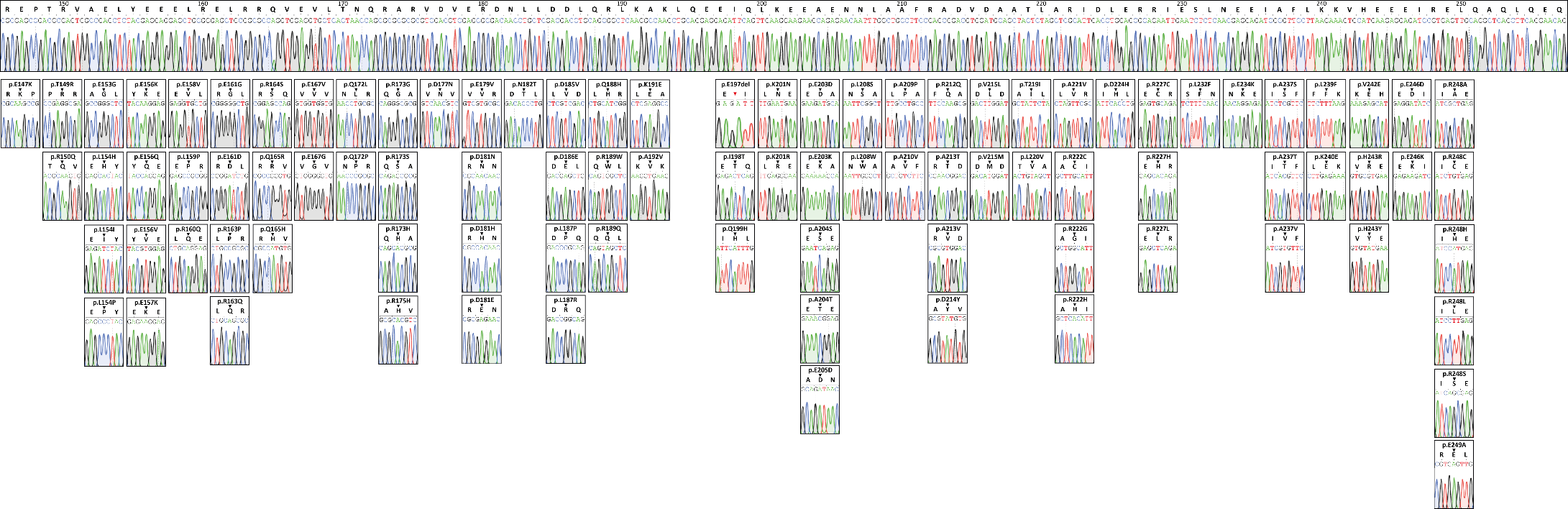

**Figure S5.** Partial electropherograms of the desmin expression constructs (pEYFP-N1-DES) carrying proline mutations at each position within the 1B domain.

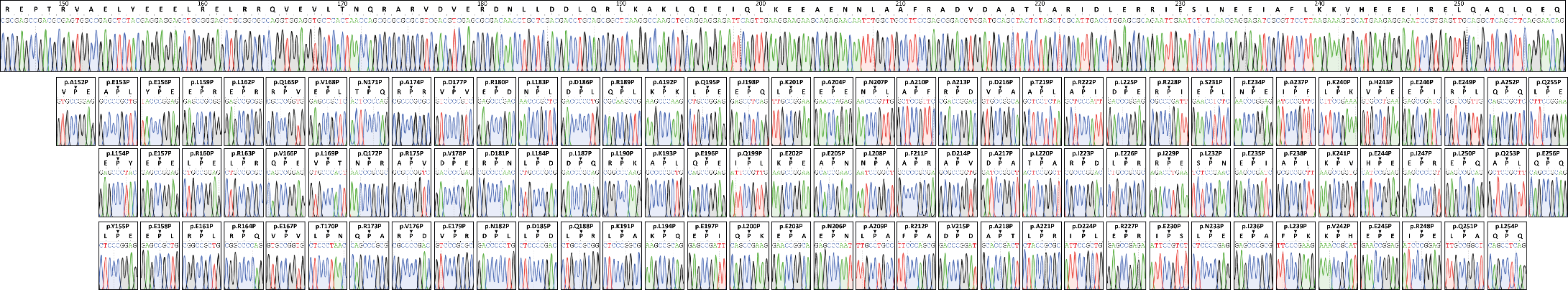

**Figure S6.** Partial electropherograms of the desmin bacterial expression constructs (pET100D-DES) carrying -p.L159P, p.R163P, p.L187P and p.E197del.

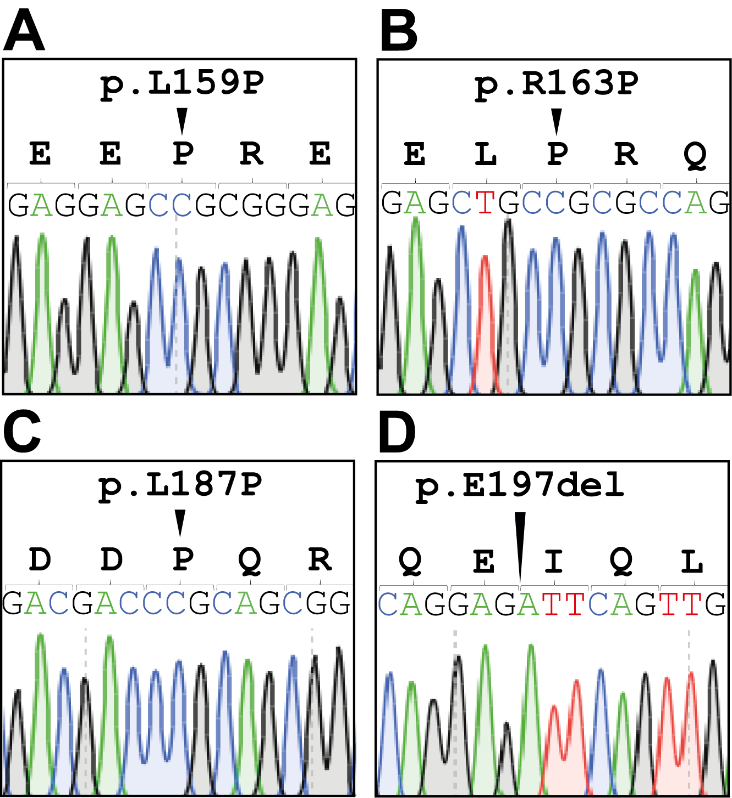

**Figure S7.** H9c2 Double transfection experiments.

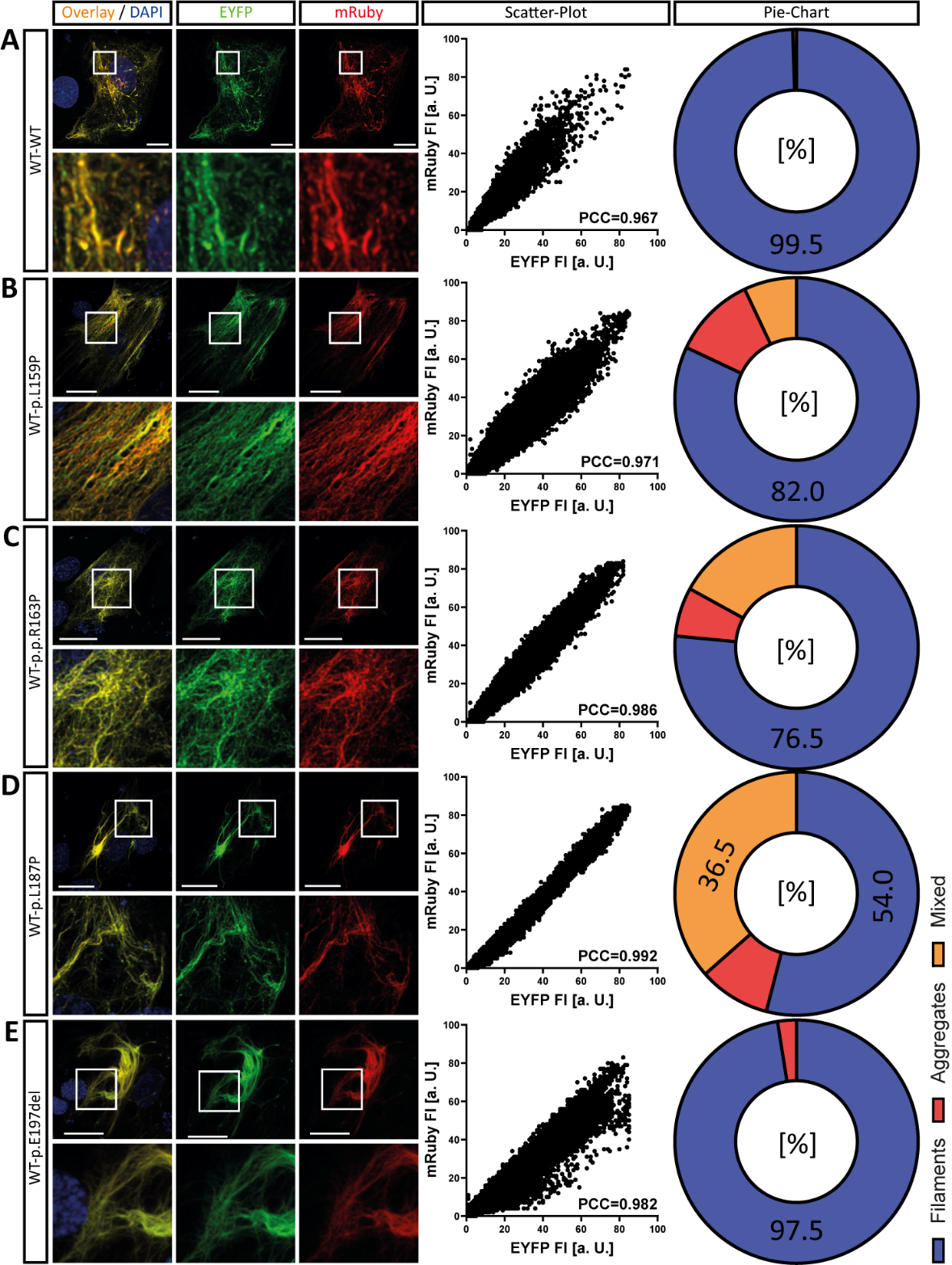

Representative images of double-transfected H9c2 cells expressing wild-type desmin (mRuby-labeled, red) and either **(A)** wild-type or **(B-E)** mutant desmin (EYFP-labeled, green) are shown. Colocalization appears as a yellow overlay, with nuclei stained in blue. Scale bars: 20 µm. Scatter plots of mRuby and EYFP signal intensities were used to calculate the Pearson correlation coefficient (PCC). The distribution of cell phenotypes - desmin filaments, aggregates, or mixed structures - is summarized in pie charts.

**Figure S8.** Decision tree for the evaluation of functional data for clinical interpretation of desmin variants according to ^1^.

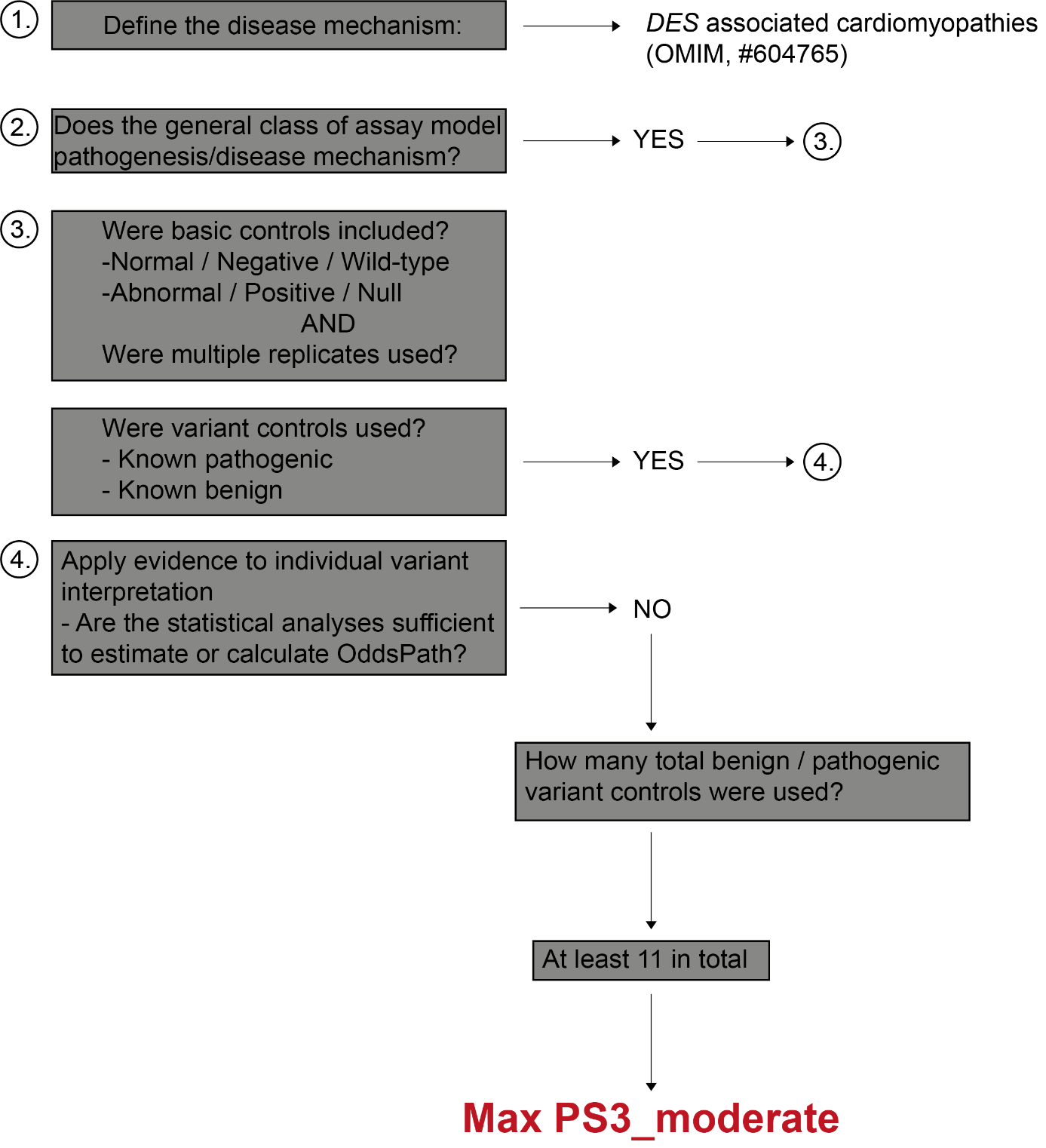
